# Assessment of Alzheimer-related Pathologies of Dementia Using Machine Learning Feature Selection

**DOI:** 10.1101/2022.04.28.22274107

**Authors:** Mohammed D Rajab, Emmanuel Jammeh, Teruka Taketa, Carol Brayne, Fiona E Matthews, Li Su, Paul G Ince, Stephen B Wharton, Dennis Wang

**Affiliations:** Sheffield Institute for Translational Neuroscience, University of Sheffield, Sheffield, UK; Department of Computer Science, University of Sheffield, Sheffield, UK; Singapore Institute Clinical Sciences, A*STAR, Singapore, 117609, Singapore; Cambridge Public Health, Cambridge, UK; Population Health Sciences Institute; Newcastle University, Newcastle upon Tyne, UK

**Keywords:** Dementia, Alzheimer’s, Feature Selection, Machine Learning, Neuropathology, Beta-amyloid

## Abstract

Although a variety of brain lesions may contribute to the pathological diagnosis of dementia, the relationship of these lesions to dementia, how they interact and how to quantify them remain uncertain. Systematically assessing neuropathological measures in relation to the cognitive and functional definitions of dementia may enable the development of better diagnostic systems and treatment targets. The objective of this study is to apply machine learning approaches for feature selection to identify key features of Alzheimer-related pathologies associated with dementia. We applied machine learning techniques for feature ranking and classification as an unbiased comparison of neuropathological features and assessment of their diagnostic performance using a cohort (n=186) from the Cognitive Function and Ageing Study (CFAS). Seven feature ranking methods using different information criteria consistently ranked 22 out of the 34 neuropathology features for importance to dementia classification. Braak neurofibrillary tangle stage, Beta-amyloid and cerebral amyloid angiopathy features were the most highly ranked, although were highly correlated with each other. The best performing dementia classifier using the top eight ranked neuropathology features achieved 79% sensitivity, 69% specificity, and 75% precision. A substantial proportion (40.4%) of dementia cases was consistently misclassified by all seven algorithms and any combination of the 22 ranked features. These results highlight the potential of using machine learning to identify key indices of plaque, tangle and cerebral amyloid angiopathy burdens that may be useful for the classification of dementia.

## Introduction

Dementia is a major healthcare concern among the elderly. It is estimated that the number of people with dementia will reach 131.5 million by 2050 worldwide [1]. There is no cure for this syndrome, but accurate and timely diagnosis of dementia may create opportunities for patients to access symptomatic and potential disease-modifying therapies. The syndrome is defined by cognitive and daily activity decline, often measured using cognitive and functional tests as defined in the Diagnostic and Statistical Manual of Mental Disorders 5th edition, along with medical history typically reported by the patient or caregiver [2]. In clinical settings further investigations are performed mostly in younger onset dementias focused on anatomical and, sometimes, functional changes measured by magnetic resonance imaging (MRI) and positron emission tomography (PET) scans, and increasingly cerebrospinal fluid (CSF) samples taken from a lumbar puncture considered to be dementia subtype biomarkers. However, dementia as it is most often manifest in older old populations is associated with multiple different pathologies [3, 4]. It remains challenging to assess the interactions of the multiple findings in the brain in relation to the syndrome expressed during life.

The Cognitive Function and Ageing Studies (MRC CFAS, CFAS I, CFAS II) are longitudinal population-based studies of ageing with a focus on cognition. This analysis focuses on the brain donation from the original MRC CFAS. More than 550 participants from CFAS voluntarily donated their brains to the study after their death for a comprehensive pathological assessment[5, 6]. Neuropathological investigations have explored the relationship of pathological features in the brain to dementia phenotypes, including various measures related to tau and beta-amyloid (Aβ) pathologies. These studies showed considerable overlap in burdens of lesions between participants dying with and without dementia [3, 4]. Dementia incidence increases dramatically as age increases and the predictive value of Alzheimer type pathologies is attenuated. Attributable risk shows the importance of many other different pathologies in the brain [7].

Machine learning (ML) classification algorithms coupled with feature selection techniques have enabled automated ways of classifying heart and skin diseases along with identifying the most informative combination of predictors of those diseases [8, 9]. Studies investigating dementia utilizing imaging assessments utilized three supervised ML algorithms (neural network, support vector machine, and adaptive neuro-fuzzy inference system (ANFIS)) for the diagnosis of Alzheimer’s disease (AD) and vascular dementia (VD) based on selected MRI features [10]. These algorithms used ranked features based on their significance in defining the class value of the dataset records. Their results show that categorizing AD and VD profiles using ML has high discriminant power with classification accuracy of more than 84% and ANFIS has a prediction rate of 77.33%. ML feature selection approaches were also shown to enable identification of neuropsychological assessments and MRI features for classification of AD [11]. Nevertheless, these ML techniques have never been used to assess the relationships between dementia and the neuropathological features of post-mortem brains from a population study.

In order to distinguish key indices such as plaque, tangle and CAA burdens, we need an objective approach to rank these pathologies and identify a combination of features useful for classifying dementia. We hypothesize that ML feature prioritisation and filtering can objectively and automatically reduce the number of neuropathological features for dementia diagnosis. To test this hypothesis, we asked several questions during the analysis of neuropathological features: 1) How are they scored across dementia cases? 2) Are any of the features related to one another and convey redundant information? 3) Can we computationally rank the features in an unbiased way to facilitate machine learning? 4) What is the smallest subset of neuropathological features needed in an ML model to explain dementia diagnosis? 5) Is there a limit to how accurately neuropathology features can classify dementia?

We investigated these questions using Alzheimer-related pathologies measured in a population-representative subcohort of CFAS [6,12–15]. There exist 34 features related to neuropathological assessments, such as Aβ features, cerebral amyloid angiopathy (CAA) features and plaque scores. These features were automatically ranked, filtered and included in ML classifiers. We also report the limits of ML classification of dementia using neuropathology factors and discuss possible reasons for these limitations.

## Material and methods

### Overview of feature selection approach

The selection of neuropathology features that are informative of dementia involved three stages of design, implementation and evaluation (Fig. 1). We obtained access to and downloaded the CFAS dataset following review and ethics approval (REC 15/WA/0035) by the CFAS management committee. Re-coding of available neuropathological features was performed to categorize and label them into distinct categories (tau, Aβ, demographics, etc.). We used supervised learning and feature selection techniques based on multiple filter-based methods. Features were then ranked according to their importance, and the most significant features identified. The smallest subset of features that can classify dementia was identified from the significant features using advanced supervised ML techniques. We determined how neuropathological features combined to detect dementia. We then identified misclassified cases using neuropathology features and linked the associations with other non-standard pathologies.

**Fig. 1.**
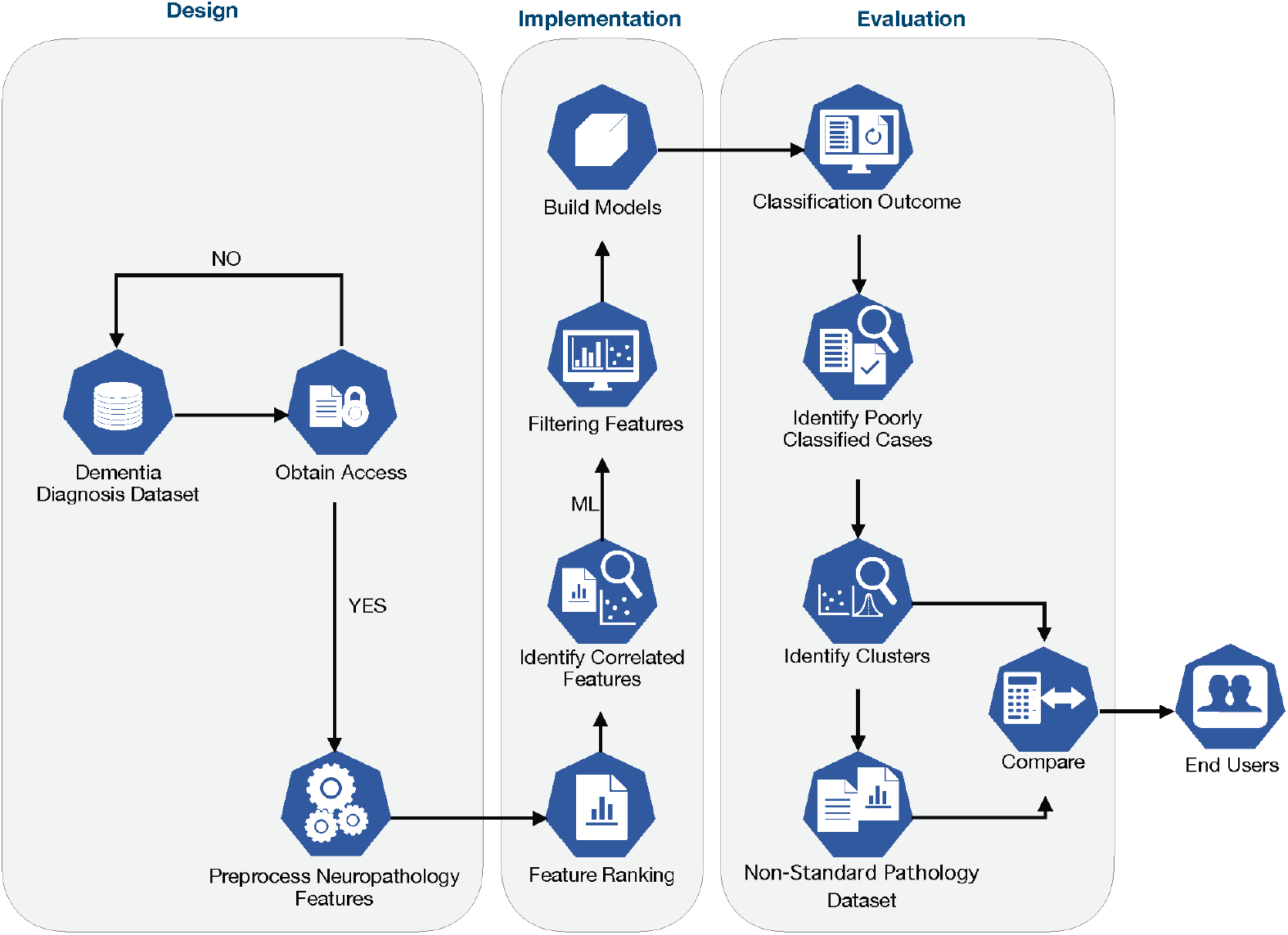
Methodology for classification of dementia. The methodology for classification of dementia followed three stages of design, implementation, and evaluation. After acquiring access to neuropathology and additional data, we pre-processed and assessed feature-feature correlation. We used feature ranking via filter methods to rank all neuropathology features. Classifiers were then benchmarked with different subsets of features selected according to their rankings. We then compared cases that were consistently misclassified, and evaluated brain attributes associated with these cases in order to improve machine learning.

### CFAS cohort

The CFAS cohort used for this study included data from two of the centres (Cambridge and Newcastle), totalling 186 subjects with 34 features in addition to age and brain weight as shown in Table 1. The features include basic neuropathological measures for each subject, including Braak neurofibrillary tangle (NFT) stage, Brain-Net Europe protocol for tau pathology, hippocampal tau NFT stage [16], Thal phase, primary age-related tauopathy (PART), cerebral amyloid angiopathy (CAA), thorn-shaped astrocytes (TSA) [14] and microinfarct stage [17] (Table 1). A total of 107 out of the 186 subjects had a diagnosis of dementia, which represents approximately 58% of the cohort. Of these 107 cases, 72 were women and 35 men, their median ages were 89 and 88. There was a balanced gender ratio (37 females and 33 males) for participants dying without dementia (median age 85, 79).

**Table 1:** description of the neuropathology features in addition to the age and brain weight features.

| No | Feature | Feature Description | Type | Control |  |  |
| --- | --- | --- | --- | --- | --- | --- |
|  |  |  |  | Dementia (n=107) | No Dementia (n=69) | Missing (n=9) |
| 1 | Braak NFT stage | Braak Stage refers to the Braak neurofibrillary tangle (NFT) stage (0-VI) [18,19]. | Nominal | 107 | 69 | 0 |
| 2 | Thal phase | Thal phase refers to the Thal Aβ phase, which is the new BrainNet stage for Aβ to detect immunopositive amyloid in cortical and subcortical areas and differentiate five phases [20,21]. | Nominal | 107 | 69 | 0 |
| 3 | Aβ stage typical | Aβ stage typical indicates the Aβ stage typical and atypical [15]. | Nominal | 107 | 69 | 0 |
| 4 | PART-definite | PART relates to the new primary age related-tauopathy concept. PARTdefinite as cases having no Aβ pathology (Thal 0) and with Braak NFT stage I-IV [22]. | Nominal | 50 | 46 | 80 (45%) |
| 5 | PART-all | Those cases with mild Aβ pathology (Thal I-II) and with Braak NFT stage I-IV [22]. | Nominal | 71 | 62 | 43 (24%) |
| 6 | CAA areas | The number of brain areas examined that have CAA (number of areas out of 9 maximum) [23]. | Numeric | 107 | 69 | 0 |
| 7 | CAA type | As defined by Thal where CAA type 1 are cases with capillary amyloid and 2 only in larger vessels and type 0 no CAA [12,23]. | Nominal | 107 | 69 | 0 |
| 8 | CAA parenchymal | CAA severity score according to Love et al [24] leptomeningeal and parenchymal vascular amyloid in four neocortical areas- So in any area CAA can be 1, 2, or 3 and the score ranges from 0 to 12 [15]. | Nominal | 107 | 69 | 0 |
| 9 | CAA meningeal | CAA severity meningeal has the same scoring system as CAA parenchymal with the score ranging from 0 to 12 [15]. | Nominal | 107 | 69 | 0 |
| 10 | CAA total severity | The scores for parenchymal and leptomeningeal amyloid were summed in four areas, scores range from 0 (minimum) to 24 (maximum) for severity in cortical areas [23]. | Numeric | 107 | 69 | 0 |
| 11 | CAA frontal | CAA in frontal cortex (Present or Absent) [16]. | Nominal | 107 | 69 | 0 |
| 12 | CAA temporal | CAA in temporal cortex (Present or Absent) [16]. | Nominal | 107 | 69 | 0 |
| 13 | CAA parietal | CAA in the parietal cortex (Present or Absent) [16]. | Nominal | 107 | 69 | 0 |
| 14 | CAA occipital | CAA in occipital cortex (Present or Absent) [16]. | Nominal | 107 | 69 | 0 |
| 15 | CAA hippocampus | CAA in hippocampus and occipitotemporal gyrus (Present or Absent) [16]. | Nominal | 107 | 69 | 0 |
| 16 | CAA cerebellum | CAA in cerebellum (Present or Absent) [16]. | Nominal | 106 | 68 | 2 (1%) |
| 17 | BrainNet tau stage | BrainNet tau stage, refers to BrainNet Europe Protocol for tau pathology, a six-stage scheme that uses neuropil threads and is proposed by the BrainNet Europe Consortium [20]. | Nominal | 107 | 68 | 1 (1%) |
| 18 | Hippocampal tau NFT stage | Hippocampal tau neurofibrillary tangles (NFT) stage | Nominal | 56 | 35 | 85 (48%) |
| 19 | subpial TSA in expanded cortex | The subpial thorn-shaped astrocytes (TSA) in the expanded cortex. | Nominal | 107 | 68 | 1 (1%) |
| 20 | subpial TSA in mesial temporal lobe | The subpial thorn-shaped astrocytes (TSA) in the mesial temporal lobe. | Nominal | 107 | 68 | 1 (1%) |
| 21 | subpial TSA in brainstem | The subpial thorn-shaped astrocytes (TSA) in the brainstem. | Nominal | 107 | 68 | 1 (1%) |
| 22 | TSA-any | Thorn-shaped astrocytes (TSA) in any brain area. (Present or Absent) | Nominal | 107 | 68 | 1 (1%) |
| 23 | TSA-total | The number of areas in the brain with thorn-shaped astrocytes (TSA) [25–28]. | Numeric | 107 | 68 | 1 (1%) |
| 24 | Tufted astrocytes | The tufted parenchymal astrocytes in any brain area. | Nominal | 107 | 68 | 1 (1%) |
| 25 | Subpial mesial temporal | The subpial tau neurites in the mesial temporal lobe. | Nominal | 107 | 68 | 1 (1%) |
| 26 | Subpial brainstem | The subpial tau neurites in the brainstem/subcortical region. | Nominal | 107 | 68 | 1 (1%) |
| 27 | Argyrophilic grains | The argyrophilic grains disease. | Nominal | 107 | 68 | 1 (1%) |
| 28 | Cortical stage | The cortical microinfarcts stage which distinguishes the number of cortical areas that have microinfarcts. | Numeric | 106 | 69 | 1 (1%) |
| 29 | Subcortical stage | Subcortical lacune stage which distinguishes the number of subcortical areas that have microinfarcts. | Numeric | 106 | 69 | 1 (1%) |
| 30 | Microinfarct stage | The total microinfarct stage which differentiate the number of total areas that have microinfarcts. | Numeric | 106 | 69 | 1 (1%) |
| 31 | Frontal microinfarct | Frontal microinfarct [17]. | Nominal | 106 | 69 | 1 (1%) |
| 32 | Temporal microinfarct | Temporal Microinfarct [17]. | Nominal | 106 | 69 | 1 (1%) |
| 33 | Parietal microinfarct | Parietal microinfarct [17]. | Nominal | 106 | 69 | 1 (1%) |
| 34 | Occipital microinfarct | Occipital Microinfarct [17]. | Nominal | 106 | 69 | 1 (1%) |
| 35 | Age | Patient's age at death. | Numeric |  |  |  |
| 36 | Brain weight | Patient's brain weight. | Numeric |  |  |  |
| 37 | Dementia Status | Class Label (Dementia or No dementia) Status of a patient | Binary | 107 | 69 | 0 |

### Diagnosing dementia

Dementia status at death for each respondent was determined based on interviews/assessments during the last years of the respondent’s life. This included using the full Geriatric Mental State-Automated Geriatric Examination for Computer Assisted Taxonomy diagnostic algorithm, the Diagnostic and Statistical Manual of Mental Disorders (third edition -revised), interviews of the informants after the respondent’s death and cause of death. Respondents were classified as having no dementia at death if they had not been identified with dementia from their last interview if less than six months before death, or if they did not have dementia identified at the last interview and the retrospective interview showed no dementia at death. Bayesian analysis was used to estimate the probability of dementia where their last interviews were more than six months before death, and no record of having dementia at interview and no retrospective informant interview (RINI) [5, 29].

### Ranking neuropathology features

To gain preliminary insight and highlight influential neuropathological features of dementia, we used different filter-based feature selection methods to measure the relevance of each feature to dementia. These included Chi-square (CHI) [30], Gain Ratio [31], Information Gain (IG) [32], ReliefF [33, 34], Symmetrical Uncertainty [35], Least Loss [36] and Variable Analysis [37, 38]. Generally, filter methods use different mathematical models to compute feature relevance. Filter-based methods are efficient feature selection methods that employ mathematical models to derive scores for each feature based on correlations between the features and class label in the input dataset. The filter methods use a different mathematical criterion to compute scores, where the scores vary based on the type of the filter method used. There can be discrepancies in the ranking of features based on such scores due to the different mathematical models used [38, 39]. The CFAS cohort consisting of 186 post-mortem and 34 neuropathology features was used for feature ranking. In addition to the 34 neuropathology features, age and brain weight (adjusted for gender) were included. Using SciPy.stats v1.5.4 in Python3, we used z-score to adjust brain weight based on sex.

CHI utilizes the difference between observed and expected frequencies of the instances as shown in Equation (1).

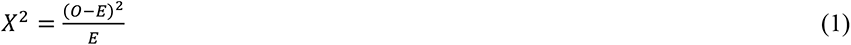

where *O* and *E* are the Observed and Expected frequencies for a specific feature, respectively. IG employs Shannon entropy to measure the correlation between a feature and dementia status (Equations 2 & 3).

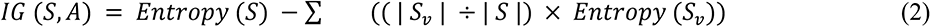

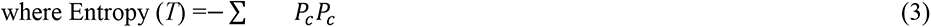

*P* is the probability that *S* belongs to class label *c*. *S_v_* is the subset of *S* for which *a* feature has value *v.* |*S_v_*| is the number of data instances in *S_v_*, and |*S*| is the size of *S*.

Gain Ratio is a normalized form of IG which is estimated by dividing the IG with the Entropy of the feature with respect to the class (Equations 4 and 5).

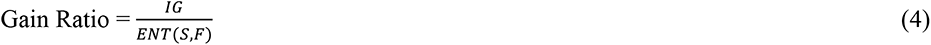

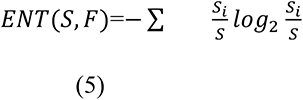

where IG denotes the information gain and *ENT* is the Entropy of feature F over a set of examples S.

Symmetrical Uncertainty deals with the bias of IG that occurs due to a large number of distinct values for the feature and presents a normalized score (Equation 6).

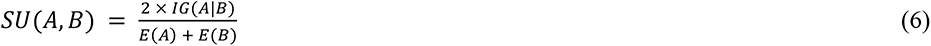

where *IG*(*A*|*B*) denotes the information gain of A after knowing the class. E(A) and E(B) are the Entropy values of A and B, respectively.

ReliefF calculates the scores of each available feature with the class using the differences between the neighbouring data instances and the target instances (Equation 7).

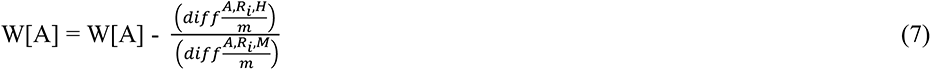

where, W[A] are the feature weights, A is the number of features, m is the number of random training data instances out of ‘n’ number of training data instances used to amend W.

*R_i_* = A random chosen test instance and H/M is nearest hit and nearest miss

Least Loss is computed per feature based on the simplified expected and observed frequencies of the features (Equation 8), and Variable Analysis employs a vector of scores of both CHI and IG results, normalizes the scores, and then computes the vector magnitude (V_score) (See Equations 9 & 10).

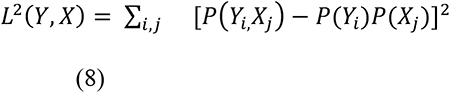

where X is the independent feature class, Y is class label, *P*(*Y_i_*) is the theoretical marginal distributions of *Y*, and *P*(*X_j_*) is the theoretical marginal distributions of X, *P*(*Y_i_*,*X_j_*) is the theoretical joint probability distributions of X and Y.

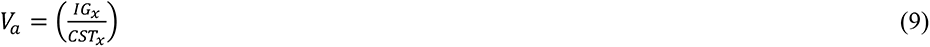

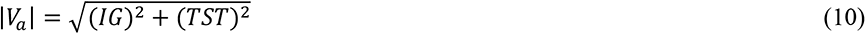

where *V_a_* is the square root of the sum of the square of its CHI and IG results of a feature.

The V_score and the Correlation Feature Set results [40] are then integrated to represent a new measure of goodness to select relevant features.

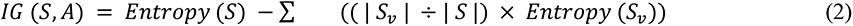

All experiment-related ranking-based feature selection was conducted using Waikato Environment for Knowledge Analysis (WEKA version 3.9.1) [41]. Percentage contribution of each feature was calculated by averaging total weights assigned by all filter methods to each feature after normalizing weights scores.

### Dementia classification

We attempted classification of dementia status in 146 out of 186 samples that had no missing values. The 146 samples had a slight imbalanced in class label with 89 demented versus 57 non-demented patients. Just before training our models, we randomly selected 57 patients once from the demented group using *sample()* function from random module in python3 and using sklearn.utils version 0.22.2.post1 to shuffle the row once, resulting in 114 samples to balance the class label and the 32 samples were held out for final assessment. We used the 114 samples during the training classifiers. The hippocampal tau stage [16] feature, which has 50% missing values, was dropped during the training classifiers. Age and brain weight were removed before training the models, ending up with 22 features and 114 samples for classification. The dataset was split into a training set of 70% (80 samples) and a testing set of 30% (34 samples).

Seven classification algorithms were trained to classify individuals’ dementia status using the 22 top ranked features. Scikit-learn version 0.22.2.post1 was used to implement and train the ML classifiers as well as measure their performance in classifying dementia. Logistic regression was implemented using *sklearn.linear_model* package with l2 penalty, regularisation parameter C set to 1, maximum number of iteration set to 2000 taken for the solvers to converge, and other parameters set to default values. Decision tree classifier was implemented using the *sklearn.tree* package. k-Nearest neighbours was implemented using the *sklearn.neighbors* with number of neighbors set to 5, the function “uniform weights” used in prediction, and “minkowski” distance metric utilized for the tree, with other parameters set to default values. Linear discriminant analysis classifier was implemented using *sklearn.discriminant_analysis* package with singular value decomposition for solver hyperparameter and other parameters set to default values. Gaussian Naïve Bayes class was implemented using *sklearn.naive_bayes*, Support Vector Machine with a Radial Basis Function kernel (SVM-RBF) was implemented using *sklearn.svm* with regularisation parameter C set to 1, the kernel coefficient gamma= “scale”, and other parameters set to default values. Support Vector Machine with linear kernel (SVM-LINEAR), was implemented using the *sklearn.svm* package with regularisation parameter C set to 1, with a “linear” kernel, gamma coefficient “scale”, and other parameters set to default. The *sklearn.metrics* package was used to report performances. Training and performance evaluation were performed 500 times from which average performance was calculated as the overall performance. Accuracy, balanced accuracy, F1 score, precision, sensitivity, and specificity utilizing regression plot, were used to determine classification performance. ML models and feature selection libraries were built using Python 3.7.3.

The smallest subset of features that can diagnose dementia with acceptable performance was determined from the 22 top ranked features. We initially created a feature set that contained the single top ranked feature *N(1)* which was used to train the ML algorithms to classify dementia, and calculate their classification performances. The second top ranked feature was added to the features subset, to generate a feature set with *N(1)+1* features. The ML classifiers were trained using the new feature subset and the classification performances were calculated. This process was repeated in descending rank order until a feature set containing all ranked features were included in the feature set. This process resulted in *22* feature sets that ranged in size from *1 to 22*, with the performance of each feature subset in classifying dementia calculated. The best subset of features was determined as a compromise between performance and size. For each feature set, the data is split into 30% test set and 70% training set.

### Evaluation of classification performance

We formulated prediction of dementia as a binary classification problem (Dementia, Control), therefore, evaluation metrics such as accuracy, f1-score, balanced accuracy, precision, specificity, and sensitivity, are used to measure the performance of the subsets of features. The following evaluation metrics were used:

● True positives (TP): Number of dementia cases that were correctly classified.

● False positives (FP): Number of healthy subjects incorrectly classified as dementia cases.

● True negatives (TN): Number of healthy subjects correctly classified.

● False negatives (FN): Number of dementia cases incorrectly classified as healthy subjects.

● Accuracy (%): The proportion of correct classifications among total classifications:

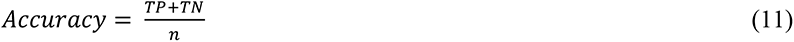

where *n* is the number of total classifications per test.

● Sensitivity (%): It is the proportion of dementia cases correctly classified.

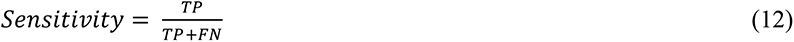

● Specificity (%): It is the proportion of healthy subjects correctly classified.

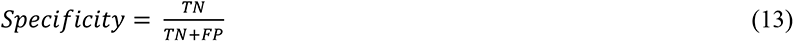

● Precision: It is the proportion of subjects classified as dementia cases who actually have dementia.

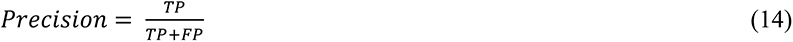

● F1 score (F-measure) (%): Harmonic mean of precision and sensitivity.

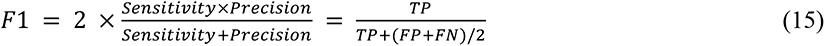

### Identifying misclassified cases

We further investigated the classification performance for all seven classifiers Logistic regression, Decision tree, k-Nearest neighbours, Linear discriminant analysis, Gaussian Naïve Bayes, SVM-RBF, and SVM-LINEAR using Scikit-learn version 0.22.2.post1 [42] in Python3 on each sample. Leave one out cross-validation was used for training and performance evaluation of trained classifiers using Scikit-learn version 0.22.2.post1 [42] in Python3. A *split()* function was provided with the dataset to enumerate for training and test sets evaluation, where evaluation was achieved by making predictions and comparing the prediction values versus the expected values. The classification algorithms trained using the top ranked 22 subset of features and 114 samples, where one feature is added at a time run creating 22 subsets of features for each classifier. All samples were clustered into true positive & true negative, false positive and false negative based on the performance of each classification run, and visualized using a heatmap to highlight the differences. The “*clustermap*” function in Seaborn package version 0.11.0 [42] was used for hierarchical clustering. The “average” was used for the linkage method in the cluster map, and “euclidean” distance was used as the distance metric. ML models and feature selection libraries were built using Python 3.7.3.

To identify pathological and demographic features (Table 2) distinguishing the three clusters of classification performance, we used robust feature selection based on Recursive Feature Elimination (RFE) with a linear SVM as the estimator [43] to identify the smallest set of non-standard pathologies features for each of the three clusters [44]. This technique has a good balance between performance and computational cost [45]. The linear SVM is initially trained using the full feature set of the training data with the C-parameter set to one. The absolute weights in the weights vector of the hyperplane is of the trained model were used to rank features according to importance, and the worst performing feature pruned from the feature set. This process is repeated until the required number of features in the signature is achieved. For a dataset with *J* samples and *K* features, M=100 subsamples are randomly sampled, and feature selection carried out in each subsample and classification performance calculated. Different sizes of signatures for each cluster, ranging from one to the full feature set. Each feature set was used to train an XGBOOST model to classify the cluster against the rest of the clusters [46]. The best signature of features for each cluster was chosen as a trade-off between signature size and classification performance. Accuracy and F1-score were used as classification metrics. ML models and feature selection libraries were built using Python 3.8.5, Scikit-learn 24.2 library and Jupyterlab 2.2.6.

Links for python script codes in GitHub, (https://github.com/mdrajab/CFAS-ranking-code) for the processes of ranking neuropathology features and classification models and (https://github.com/emmanueljammeh/cfas) for feature signatures showing association of the non-standard pathologies and demographics features with clusters.

## Results

### Distribution of neuropathology feature scores across dementia cases

Fig. 2 depicts the distribution of values of participants dying without and without dementia across all neuropathology features in our study containing 186 samples and 34 attributes. In addition to the 34 neuropathology features, age and brain weight (adjusted for gender) features were included. People aged between 80 and 89 years had a higher frequency of dementia than other age sub-groups. The findings from this analysis are those already published using more traditional biostatistical methods, with the proportion of individuals with dementia increasing with increasing Braak NFT stage, peaking at stage IV, with the same finding for Thal phase increase, hippocampal tau stage. The measures of CAA across subjects reveals that the proportion of dementia cases increases as the number of brain areas with CAA increases. On the other hand, few microinfarcts features (frontal, occipital, parietal) were observed in individuals who died with dementia. A similar observation is seen with Aβ stage typical and Argyrophilic grains, which may limit classifiers from differentiating subjects using these features.

**Fig. 2.**
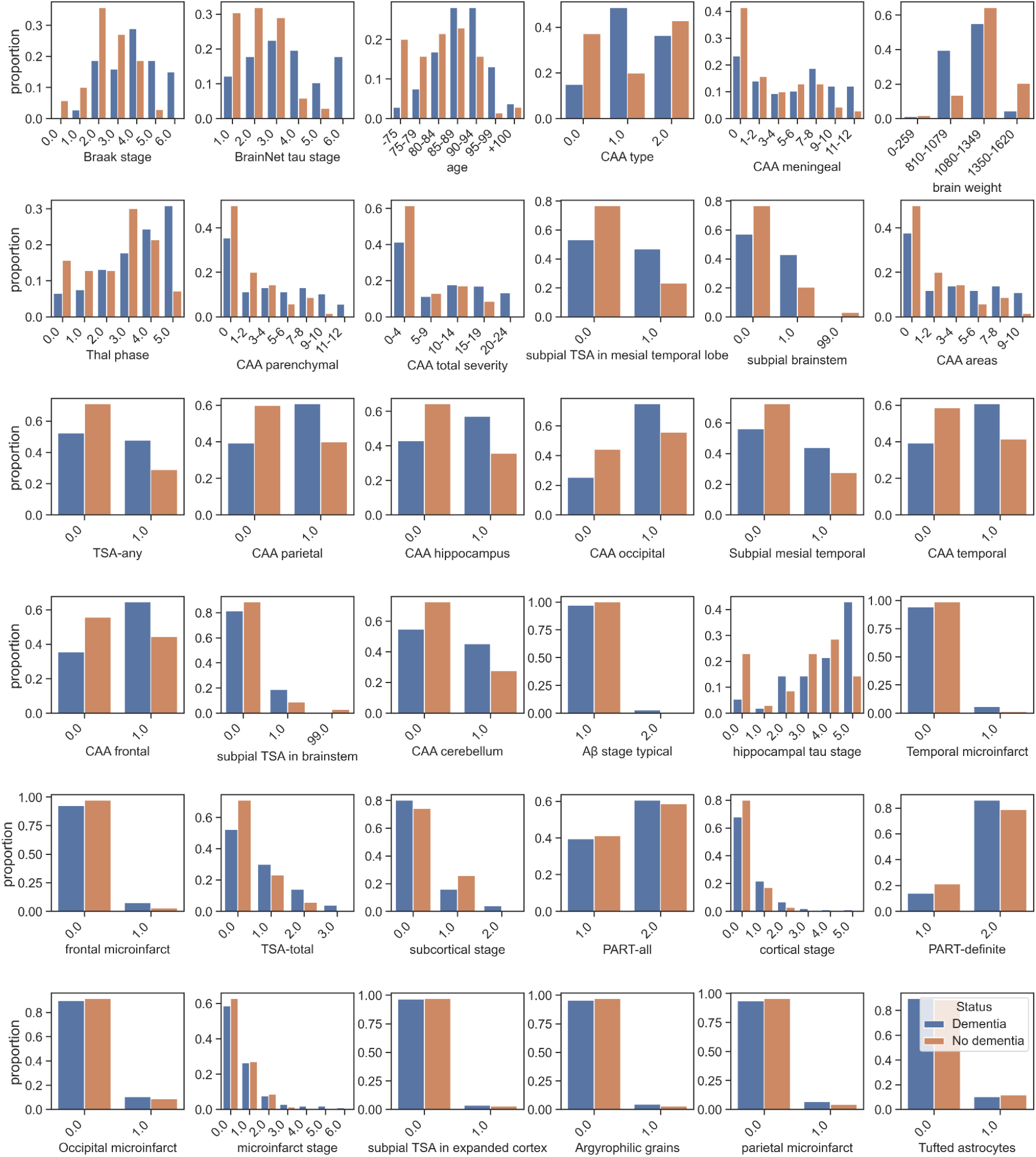
CFAS Neuropathology features distribution. The figure depicts neuropathology features distribution including age and brain weight (proportion of individuals with and without dementia of the CFAS neuropathology Dataset). All features shown are based on the ranking features list, row from left to right. Most features are categorical features except some are numeric, such as age, CAA total severity, brain weight, CAA areas, TSA-total, cortical stage, subcortical stage and Microinfarct Stage.

### Highly correlated neuropathology features

The results of statistical analysis to determine the significance of each feature and to determine the inter correlation of features identified some highly correlated features such as CAA related features. Since CAA-related features including CAA type, CAA areas, and CAA total severity (CAA meningeal, CAA parenchymal) are common among the top features offered by the different feature selection results shown (Supplementary Table 1, Additional File 1), we needed to ensure that only dissimilar features are chosen by minimizing feature-to-feature correlations. We identified three main clusters of highly correlated features as shown in (Fig. 3) using all neuropathology features in our study containing 186 samples and 34 attributes in addition to age and brain weight features. Hence, some of these features may be redundant in the diagnosis of dementia based on neuropathological features.

**Fig. 3.**
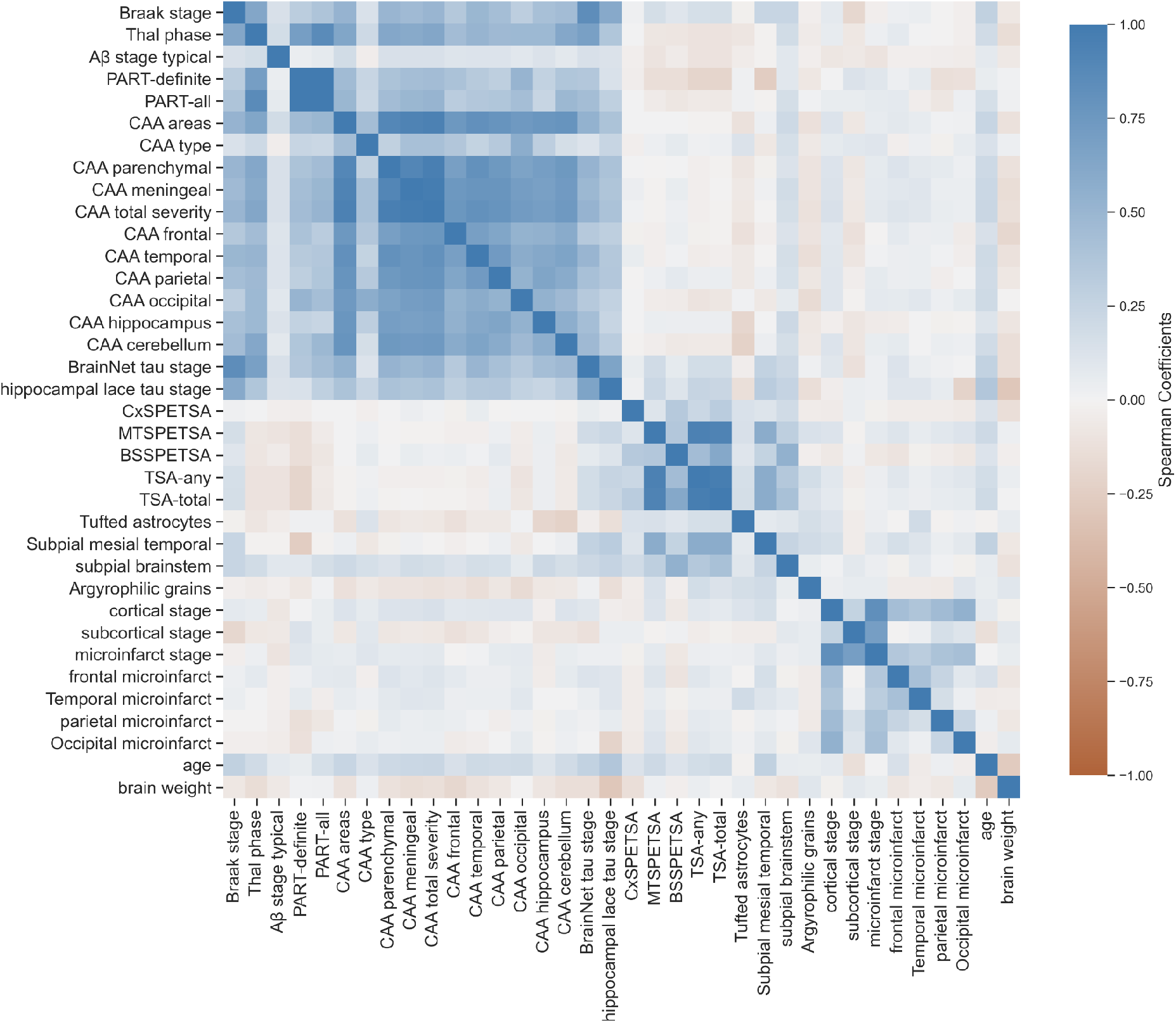
Spearman correlation of the complete CFAS neuropathological data set. Heat map Spearman correlation of the complete CFAS neuropathological data set 34 neuropathology features in addition to age and brain weight features as a benchmark, 36 features in total and 186 samples. A coefficient close to 1(blue colour) means that there is a very high positive correlation between the two variables. The diagonal line is the same variable i.e. spearman rho 1.

### Ranking of neuropathology features

The ranking results of statistical tests conducted to determine the significance of each feature to dementia using seven feature-ranking methods are presented in (Supplementary Table 1, Additional File 1). The results show that a few common features are highly ranked by the seven different ranking methods. Braak NFT stage, BrainNet tau stage and CAA-related features are consistently ranked at the top by most of the feature selection methods. A high ranking of Braak NFT stage which shows the neurofibrillary tangle stage (0–VI), supports Braak NFT stage as a highly impactful feature for dementia pathology [19]. The fact that the Braak NFT score was ranked in the top 6 by all ranking techniques (CHI, gain ratio, information gain, ReliefF, symmetrical uncertainty, least loss and variable analysis) is important to both human and computer-aided diagnosis of dementia, and that it should be utilized as a primary feature. Different feature selection techniques result in different ranking of the features, however, most of the commonly used features are consistently highly ranked. For example, Braak stage, BrainNet tau stage, CAA type, Thal phase, subpial brainstem, and subpial TSA in mesial temporal lobe are consistently ranked in top 12 (out of 36) notwithstanding which ranking method is used. Nevertheless, averaging feature ranks across different ranking methods should lead to a more accurate ranking of the features in CFAS dataset.

BrainNet tau stage appears at the top of ranked features and it has previously been found to be highly correlated with the Braak NFT stage as tangles and neuropil threads seem to progress together [14]. BrainNet tau stage, a six-stage scheme that uses neuropil threads and proposed by the BrainNet Europe consortium [47], has been used to predict dementia pathology in recent research studies. CAA-related features including CAA type, CAA areas, and CAA total severity (CAA meningeal, CAA parenchymal) are common among the top features offered by the different feature selection results shown in (Supplementary Table 1, Additional File 1). We believe this may be in part due to the high correlation among these CAA related features (Fig. 3). Therefore, we evaluated these features to ensure that only dissimilar features are chosen by minimizing feature-to-feature correlations. Lastly, subpial TSA in mesial temporal lobe appears frequently in the results of all feature selection methods with a high rank. This indicates that tau-related factors such as subpial TSA in mesial temporal lobe can quantify dementia neuropathological factors especially when they present in the aged human medial temporal lobe.

All 34 neuropathology features in addition to age and brain weight (gender adjusted) and 186 samples were assessed using the seven ranking methods (Supplementary Table 1; Figur 4, Additional File 1). Based on the weights from each ranker we calculated the percentage of contribution for each feature by taking the average of total weight assigned by all filter methods to each feature. We found a subset of 25 features where a percentage of contribution was estimated by all ranking methods. For downstream analysis, we dropped features with less than 7% contribution or those not ranked by all ranking methods. In order to assess the utility of neuropathology features to classify dementia, we removed the non-neuropathology features (age and brain weight), and hippocampal tau stage due to high missingness, leaving 22 ranked features.

### Classification of the ranked neuropathology features

We further investigated subsets of the top 22 ranked neuropathological features and 114 samples using ML feature selection based on the results presented in (Fig. 4). A single feature was successively added from the set of 22 most significant features to create subsets of features with sizes ranging from 1 to 22 (from top to last ranked features). A sub dataset was then created for each subset of features. Each sub dataset was randomly split into a training set containing 70% of the samples and the remaining 30% of the samples were used for testing. The training set was used to train classification models using logistic regression, decision tree, k-nearest neighbours, linear discriminant analysis, gaussian naïve bayes, SVM-RBF, and SVM-LINEAR classification algorithms. The performance of each trained model was evaluated using the test set for prediction. (Fig. 5) depicts F1-score performance of all subsets of features (by forward and backward order of ranked features) in classifying dementia status for the seven ML techniques considered. The F1 score shows that top eight features have the highest performance of 74% by (SVM-RBF and logistic regression) algorithms. This supports the accuracy and balanced accuracy that show the top eight features performance of 74% for both accuracy and balanced accuracy by most algorithms. The results showed that there was no substantial classification performance increase beyond using eight features. In fact, increasing the number of features beyond 8 features resulted in a slight drop in the performance in most of the trained models in identifying dementia patients. The results suggest that for the data used in this study, the most influential neuropathology features for classifying dementia status are the eight top ranked features as shown in (Fig. 4) where age and brain weight features are excluded. We also showed sensitivity and specificity for all of our models to explain why some of our training of the forward ranking performances increase when adding the last three features (Supplementary Figures 3 & 4, Additional File 1). This explains that some features have an imbalanced class where last features in our forward ranking training show high specificity and low sensitivity. For example, in the linear discriminant analysis classifier, the last five features achieved 84% sensitivity and 50% specificity, which explained that the target class is still imbalanced for those features even though we balanced the target class for the dataset. The highest sensitivity was achieved with logistic regression 79% with top eight features, and SVM-LINEAR 79% with top ten features. For specificity, the highest specificity was achieved by a decision tree with top two features.

**Fig. 4.**
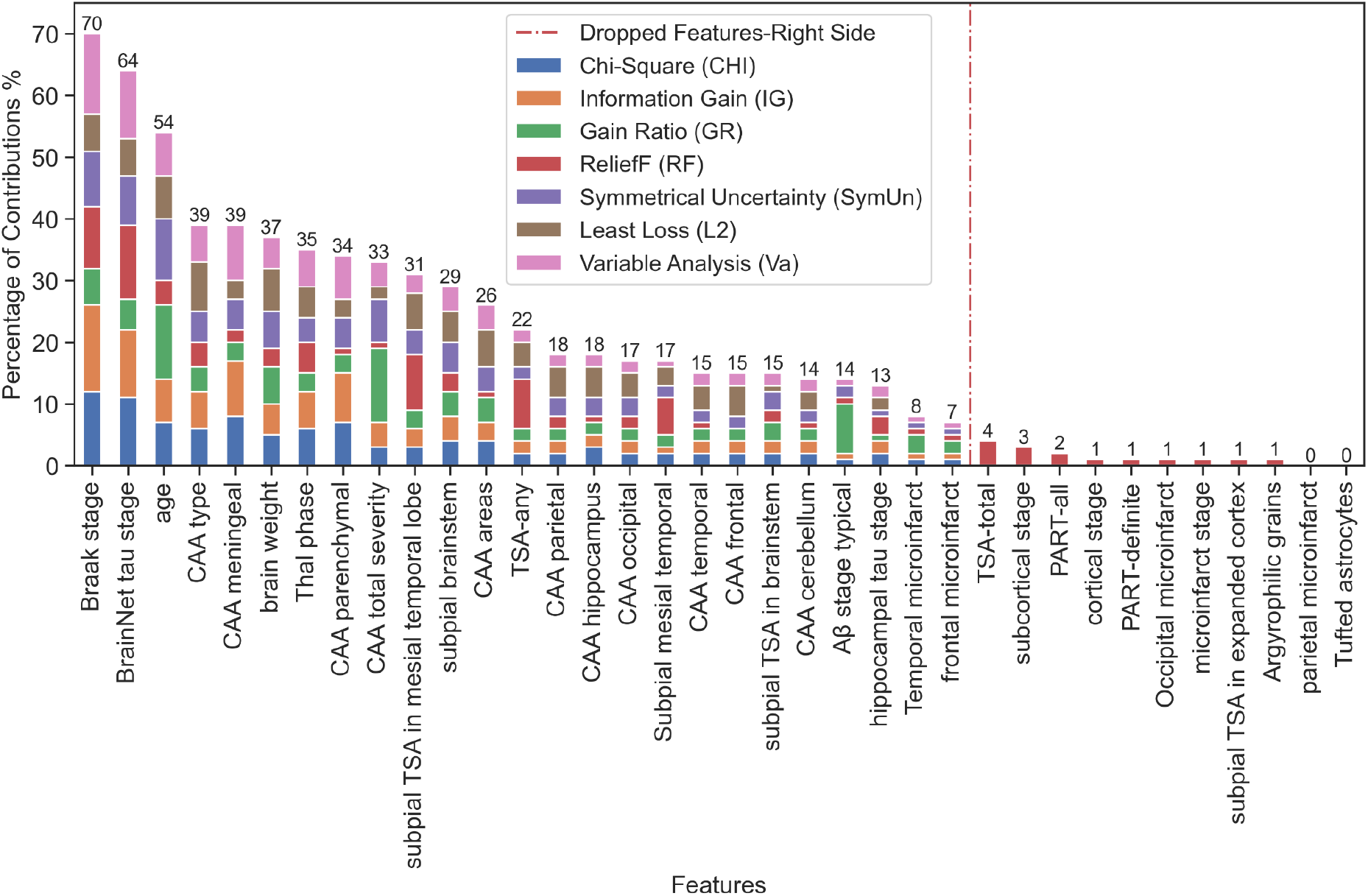
Ranking of neuropathology features. Ranking of 34 neuropathology features using seven filter methods in addition to age and brain weight features as a benchmark, 36 features in total and 186 samples. Based on these weights we calculated the percentage of contribution for each feature by taking the average. Dotted line indicates features to be dropped, which features percentage contribution show less than 7%.

**Fig. 5.**
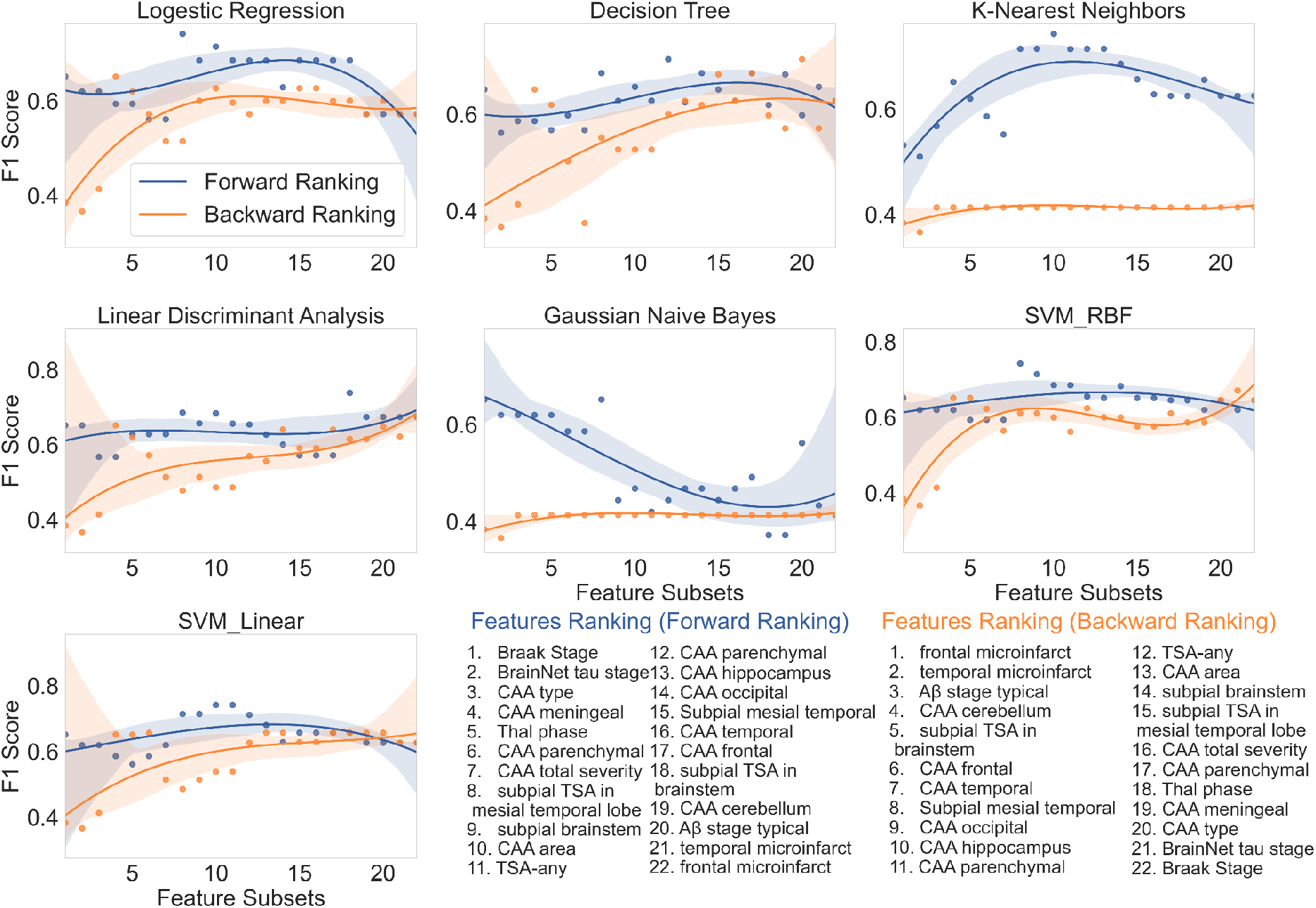
Performance of all subsets of neuropathology features. F1 score performance of all subsets of neuropathology features from the rank list forward and backward rankings. Forward ranking (blue) adds to the classifier model from the top feature to the lowest feature while the backward ranking (orange) adds to the model from the lowest feature to the top feature. Seven classifiers were utilized in this investigation: logistic regression, decision tree, k-nearest neighbors, linear discriminant analysis, gaussian naive bayes, support vector machines with radial basis function kernel, and support vector machines with linear kernel. Please see (Supplementary: Figures [1–7], Additional File 1) for other metrics such as accuracy, balanced accuracy, sensitivity, specificity.

### Limits to the accuracy of classification of neuropathology features

Classification results of different feature subsets using the seven classifiers, 114 samples and 22 top ranked neuropathology features showed that 40.4% of patients were misclassified out of 114 individuals using cross-validation. The cause of this high misclassification rate was further investigated. Using a heatmap to visualise the classification of each patient, showed that some cases are always or mostly misclassified as false positive, and false negative irrespective of the ML algorithm used. (Fig. 6) shows a heatmap of clustering the classification results, where seven classification techniques were used for the clustering of classification of patients for different numbers of features. Three clusters were identified containing cases that were classified correctly, and misclassified as false positive or false negative. The false positive cluster denotes cases where neuropathology features classified them as having had dementia when in actuality they did not. The false negative cluster denotes cases classified as not having dementia, but in actuality they did. Perhaps, this cluster could correspond to cases of dementia with insufficient neuropathology changes [48].

**Fig. 6.**
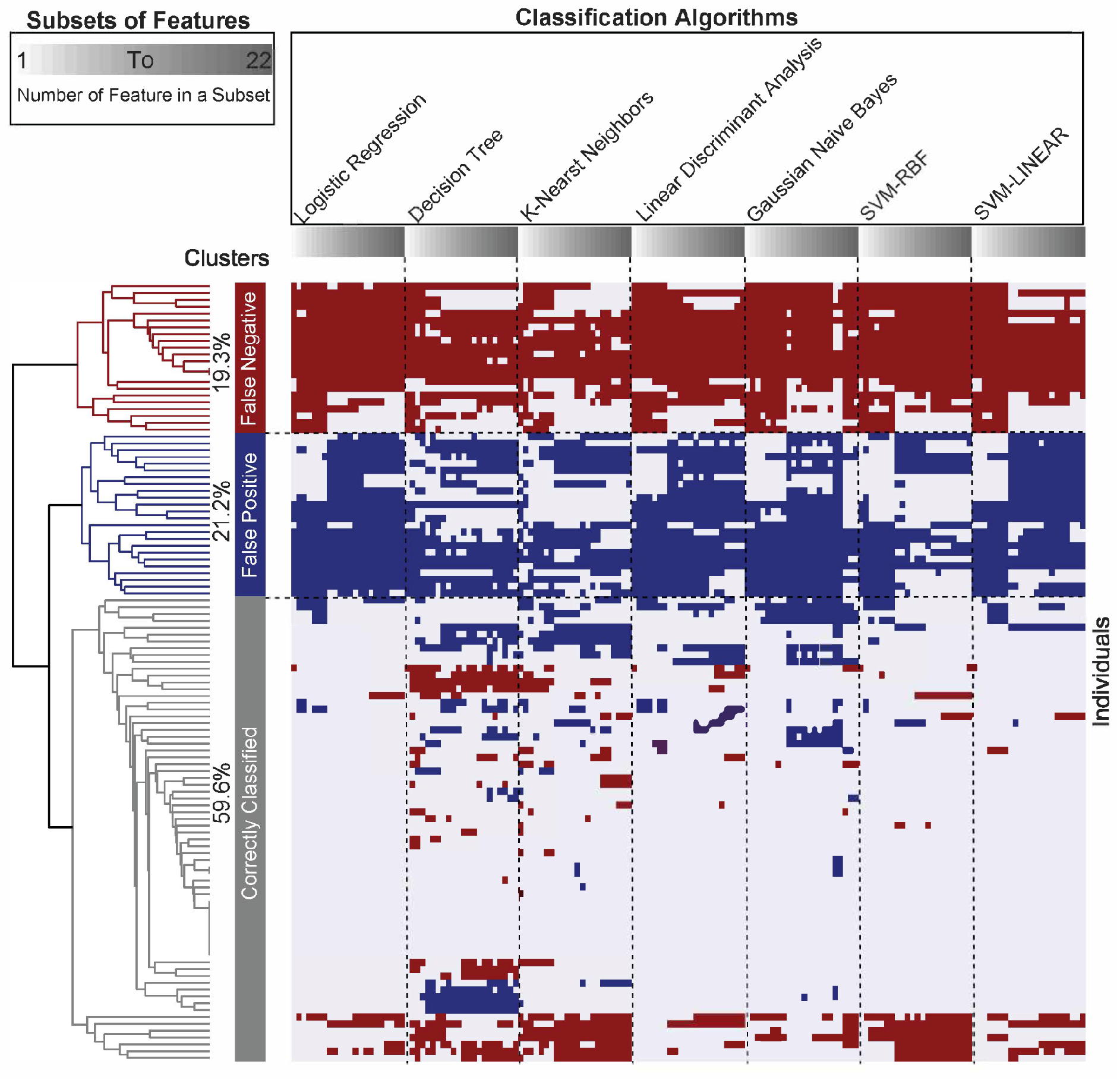
Clustering of classification performance. Clustering of classification performance from leave one out cross-validation on 114 CFAS participants and top 22 ranked standard neuropathology features. Each cluster illustrates a classification that was given to individuals consistently, or nearly consistently, irrespective of what classification algorithm was used. Evaluation of 7 classifiers revealed 24 individuals (blue) were mostly misclassified as false positive, 22 individuals (red) were mostly misclassified as false negative, and 68 individuals (grey) were mostly correctly classified as true positive or true negative. Each algorithm evaluated subsets of ranked features from 1 (top feature) to 22 features (all ranked features).

Since the classical markers of neuropathology features summarising the prevalence of plaques and tangles could not classify a large proportion of patients, we hypothesised that non-standard pathologies for rarer dementia syndromes and regional markers could be more useful. These less common and ‘disregarded’ pathologies have been described across the CFAS cohort [49]. We therefore performed further analysis to determine which features are associated with cases where the ranked neuropathology features alone were unable to explain dementia. There are 28 features used and 114 samples. The non-standard features used were based on more granular neuropathology features in different regions in the brain such as, neuronal loss, gliosis, pick bodies, lewy bodies, spongiform changes, superficial gliosis, tangles, virchow-robin space expansion and ballooned neurons as well as some demographic features such as, gender, age, and brain weight features.

Our best performing model, RFE with a SVM estimator (SVM-RFE), has been shown to be effective in the removal of features that are irrelevant and redundant to achieve good generalisation. (Fig. 7) shows the set of non-standard pathologies features measured in the different regions of the brain along with age, sex and brain weight and their coefficient values compared to the classification performance from the ML models that used standard neuropathologies (Supplementary Figure 6, Additional File 1). We found that the mean age for false negative cases was highest with mean 89.3 years, whereas the false positive mean age is 84.5 and true positive & true negative mean age is 84.7. We also found that brain weight mean was lower in the false negative group than false positive and true positive & true negative groups. Lewy bodies in substantia nigra, neuronal loss in the hippocampus, neuronal loss in substantia nigra, tangles in temporal lobe, parenchymal CAA in the frontal lobe, and gliosis in hippocampus were all associated with the classification performance of standard neuropathologies, however, a high proportion of misclassifications occurred where there was a lack of pathology detected from these measures.

**Fig. 7.**
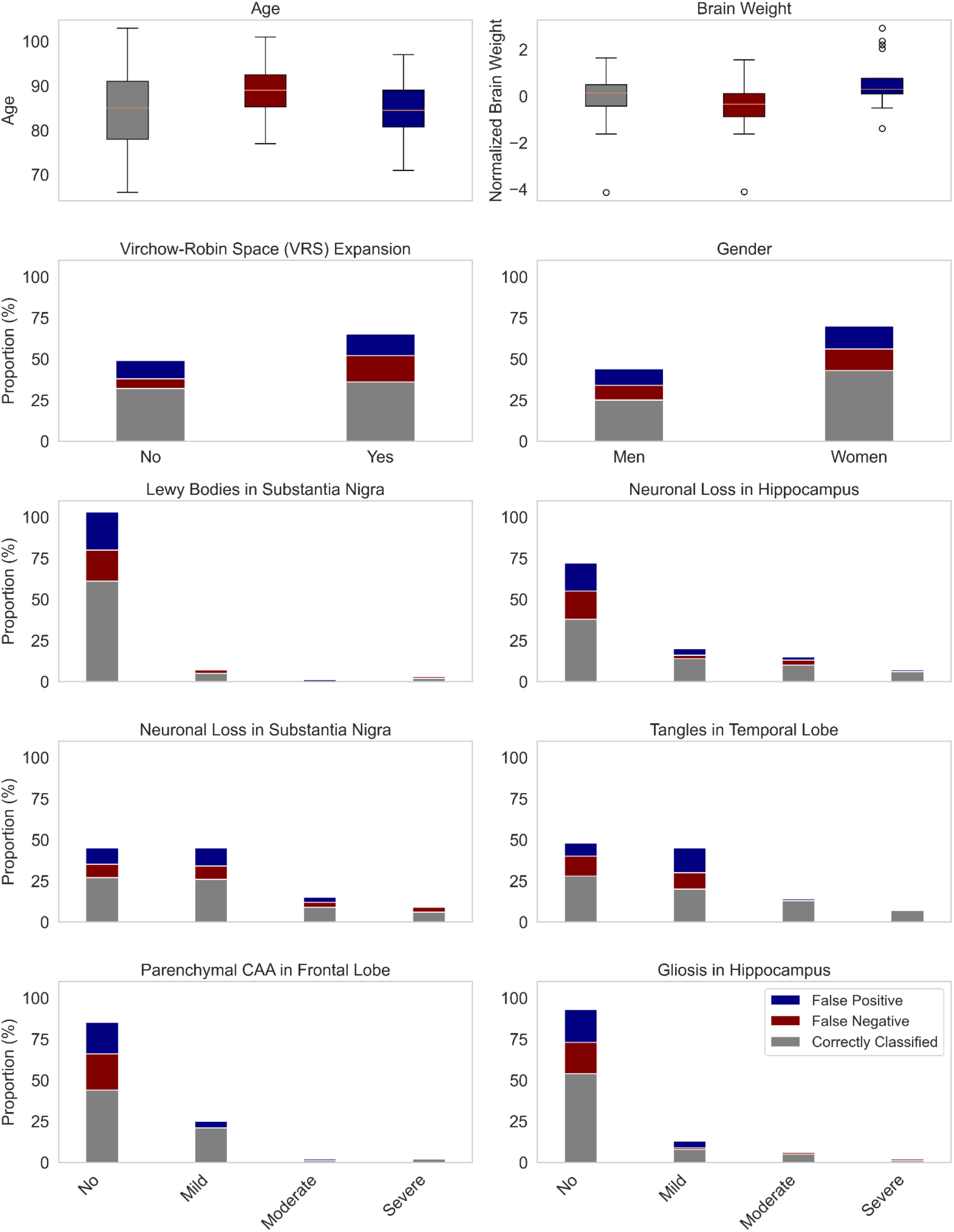
Associations of standard and non-standard neuropathological and demographic features. Non-standard neuropathological and demographic features that were associated with mis-classified and correctly classified cases by the standard neuropathology features. The features together were identified as being associated with the classification performance (Supplementary Figure 6, Additional File 1).

For further evaluation, we combined the top eight classical neuropathology features with the ten non-standard features associated with classifier performance. Together, we tested subsets of the 18 features to classify dementia status. Earlier using the classical features, we observed 40.4% of the cases being misclassified, however, when combining the feature sets the percentage of misclassified cases decreased to 35.1% (Supplementary Figure 5, Additional File 1). This decrease in misclassifications was observed in cases of at least 85 years old (46.3% to 40.3%) and observed in cases younger than 85 years (31.9% to 27.7%). Of the 32 cases that were held-out, we observed sensitivity of 68.8% (logistic regression) using the top eight neuropathology features, whereas the combined set of standard and non-standard neuropathology features achieved a better sensitivity of 81.3%.

## Discussions & Conclusions

In this study, we introduced an ML approach to describe how neuropathological features at the end of life relate to functional and neuropsychological assessments Our step- wise ML approach to rank and select Alzheimer-related pathologies for classifying dementia allowed us to investigate how the different measures, such as those related to Aβ-related assessments and tau can inform about dementia status. We ranked 22 features for ML classification from 186 CFAS subjects and found that tau-related assessments, such as the Braak NFT stage, were most influential in classifying dementia in a population-representative cohort. Seven different classification algorithms tested using different subsets of ranked features revealed that maximum classification accuracy was at most 74% with the top eight ranked features. Two groups of subjects were identified after classification (false positives, false negatives) accounting for 40.4% of all participants who were consistently misclassified no matter the classification algorithms used. In order to improve classification accuracy, we considered whether non-standard and more granular neuropathology features for particular regions of the brain could help with classification. False positives and false negatives for dementia classification were more likely for different ages and brain weights [4]. Savva et al. showed that the association between AD pathology and dementia is different between younger and older ages [7]. We found that lewy bodies in substantia nigra, neuronal loss in the hippocampus, neuronal loss in substantia nigra, tangles in temporal lobe, parenchymal CAA in the frontal lobe, and gliosis in hippocampus could complement standard neuropathology features for classifying dementia. Most of the misclassifications in our analysis occurred where there was a lack of pathology observed from these measures. This is consistent with Corrada et al. who reported that 22% of demented participants did not have sufficient pathology to account for cognitive loss [50]. Using the Vantaa 85+ cohort, Hall et al. showed that cognition and education predicted dementia but not AD or amyloid-related pathologies for the oldest old [51]. When combining the top eight neuropathology features with the non-standard pathologies’ features, we were able to improve the dementia classification for the older individuals (85 years old and above).

Statistical and ML analysis of the dataset revealed at least three clusters of highly related features. This shows that some measures in the dataset are redundant because they are so highly correlated. Removing some of the redundant features in cohort datasets such as the CFAS dataset reduces collinearity, improves the performance of feature selection and classification accuracy [52–56]. We showed that different ranking methods resulted in slightly different ranking of the features in terms of their association with dementia status. Averaging feature ranks across different ranking methods showed a more consistent and robust feature ranking. This suggests that ML-based feature ranking should be averaged across different ranking techniques [57–59]. We utilised feature selection and ML classification techniques and seven different filter-methods to rank neuropathology features. The results showed that most of the commonly used features are consistently highly ranked. Braak NFT stage and BrainNet tau stage are the top two selected features in line with previous studies [6,14,15,60,61]. However, our results also showed that subpial TSA in mesial temporal lobe had a significant rank, presenting a contradictory finding from prior studies [6]. Our results show that most of our models with top eight features obtained an accuracy of 74%, with considerable misclassification with false negatives and false positives.

One of the limitations of this study is that it does not cover a vast number of classification algorithms, especially ones that utilize deep learning, due to its relatively small sample size. Further investigation using other ML techniques if the number of cases and controls in our cohort increases. Furthermore, including more instances and possibly more neuropathological variables would be advantageous to evaluate the performance of the classification algorithms used in this study. These limitations can potentially be research directions, especially in designing and implementing an accurate and cost-effective computer-aided artificial intelligence-based diagnosis system for dementia classification. This can be done in an additional study using an independent dataset, such as the Alzheimer’s Disease Neuroimaging Initiative at (http://www.adni-info.org/) [62] or the Rush Memory and Ageing Project [63]. The challenge of using neuropathology assessments is that diagnosis of dementia has a time-lapse between the last clinical assessment and the post-mortem brain, which could be a possible reason for the classification performance’s upper limit. However, the reported dementia status also included information from those who knew the individual at the time and up to death. Another explanation for the poor classification performance is that some cases express dementia during life without classical neuropathological changes [48]. Pathological features are very different between different types of dementia such as AD, Frontotemporal Dementia, VD, Lewy Body Dementia [64–66] and their mixed occurrence is still poorly understood, despite consistent reports from neuropathological collections associated with older populations and volunteer cohorts. In order to define those misclassified clusters, we may need to investigate younger populations that exhibit separate and distinct pathologies that can be learned by classification models. The features we used are mostly confined to measures of tau and Aβ. There is a need for more extensive modelling to quantify measures of other key age-related brain pathologies, particularly vascular disease, synuclein staging and age-related Transactive response DNA-binding protein 43 (TDP43) pathology (limbic predominant age-related TDP43 encephalopathy). This would allow us to link pathology with other symptoms relating to dementia. Rather than assess associations between one feature and outcome at a time, it would be useful to investigate whether the combinations of features are associated with multiple clinical phenotypes [67–71]. This would have translational value in helping to make complex diagnoses that involve multiple clinical assessments. The feature ranking and filtering approaches could also be extended to a diverse set of data features from pathology, imaging, CSF or blood-based biomarkers.

This study provides a new approach for understanding how much cognitive and functional classification of dementia can be explained by pathological features of the brain. The application of ML as a means of robust evaluation of neuropathological assessments and scores for 186 subjects and 34 neuropathology features from the CFAS cohort, highlights the benefits and limitations of automated methods for dementia classification and points out the key indices of Alzheimer-related pathologies. While we found that as many as 22 neuropathology features could be useful for classification of dementia, tau-related assessments, such as Braak NFT stage, were most influential in the ML classifiers. Our finding that 40.4% of cases are consistently misclassified with existing features opens the door to further neuropathological assessments and more complex models for explaining dementia. We hope that further applications of this approach can lead to identifying biomarkers of early diagnosis, and improve disease management plans for patients and their families.

## Data Availability

All data produced in the present study are available upon reasonable request to the authors.
All data produced in the present work are contained in the manuscript.
All data produced are available online at
CFAS (http://www.cfas.ac.uk/cfas-i/data/#cfasi-data-request), under the custodianship of Fiona E Matthews and Carol Brayne.

## Acknowledgements

We would like to acknowledge the essential contribution of the liaison officers, the medical practitioners, their staff, and nursing and residential home staff who have contributed their time and effort in collecting data as part of the Cognitive Function and Ageing Study Group. We are grateful to our respondents and their families for their generous gift to medical research, which has made this study possible.

## Author information

### Contributions

Study design and assessment of tissue sections; SBW, PGI. Data analysis; MR, EJ, TT, DW. Writing of first draft MR, EJ, TT, DW. Data oversight and analysis results interpretation; LS, FM, CB. Contribution to interpretation and to the final manuscript; all authors. All authors read and approved the final manuscript.

## Ethics declaration

### Ethics approval and consent to participate

This study was ethically approved by the CFAS management committee for analysis to be carried out at the University of Sheffield, University of Cambridge and Newcastle University. Study title “Cognitive Function and Ageing Study brain donation cohort bioresource and fieldwork activity”. REC reference (15/WA/0035). Amendment date: 28 July 2017. IRAS project ID (147624).

### Consent for publication

Not applicable.

### Availability of data and material

Data from the CFAS study is accessible via application to the CFAS (http://www.cfas.ac.uk/cfas-i/data/#cfasi-data-request), under the custodianship of FM and CB.

## References

1. M. Prince, A. Wimo, M. Guerchet, G.C. Ali, Y.T. Wu, M. Prina, Alzheimer’s disease international (2015). world alzheimer report 2015: The global impact of dementia: An analysis of prevalence, incidence, cost and trends, Alzheimer’s Disease International, London. [Google Scholar]. (2018).

2. American Psychiatric Association, Diagnostic and Statistical Manual of Mental Disorders (DSM- 5®), American Psychiatric Pub, 2013.

3. Lancet, Pathological correlates of late-onset dementia in a multicentre, community-based population in England and Wales, The Lancet. 357 (2001) 169–175. https://doi.org/10.1016/s0140-6736(00)03589-3.

4. F.E. Matthews, C. Brayne, J. Lowe, I. McKeith, S.B. Wharton, P. Ince, Epidemiological pathology of dementia: attributable-risks at death in the Medical Research Council Cognitive Function and Ageing Study, PLoS Med. 6 (2009) e1000180.

5. C. Brayne, J. Nickson, C. McCracken, C. Gill, A.L. Johnson, Cognitive function and dementia in six areas of England and Wales: the distribution of MMSE and prevalence of GMS organicity level in the MRC CFA Study, Psychol. Med. 28 (1998) 319–335.

6. S.B. Wharton, C. Brayne, G.M. Savva, F.E. Matthews, G. Forster, J. Simpson, G. Lace, P.G. Ince, Medical Research Council Cognitive Function and Aging Study, Epidemiological neuropathology: the MRC Cognitive Function and Aging Study experience, J. Alzheimers. Dis. 25 (2011) 359–372.

7. G.M. Savva, S.B. Wharton, P.G. Ince, G. Forster, F.E. Matthews, C. Brayne, Age, neuropathology, and dementia, N. Engl. J. Med. 360 (2009) 2302–2309.

8. S. Shilaskar, A. Ghatol, Feature selection for medical diagnosis : Evaluation for cardiovascular diseases, Expert Syst. Appl. 40 (2013) 4146–4153.

9. A.K. Verma, S. Pal, S. Kumar, Prediction of Skin Disease Using Ensemble Data Mining Techniques and Feature Selection Method—a Comparative Study, Appl. Biochem. Biotechnol. 190 (2020) 341–359.

10. G. Castellazzi, M.G. Cuzzoni, M. Cotta Ramusino, D. Martinelli, F. Denaro, A. Ricciardi, P. Vitali, N. Anzalone, S. Bernini, F. Palesi, E. Sinforiani, A. Costa, G. Micieli, E. D’Angelo, G. Magenes, C.A.M. Gandini Wheeler-Kingshott, A Machine Learning Approach for the Differential Diagnosis of Alzheimer and Vascular Dementia Fed by MRI Selected Features, Front. Neuroinform. 14 (2020) 25.

11. S. Thapa, P. Singh, D.K. Jain, N. Bharill, A. Gupta, M. Prasad, Data-driven approach based on feature selection technique for early diagnosis of Alzheimer’s disease, in: 2020 International Joint Conference on Neural Networks (IJCNN), IEEE, 2020: pp. 1–8.

12. D.R. Thal, U. Rüb, M. Orantes, H. Braak, Phases of Aβ-deposition in the human brain and its relevance for the development of AD, Neurology. 58 (2002) 1791–1800.

13. M.E. Murray, V.J. Lowe, N.R. Graff-Radford, A.M. Liesinger, A. Cannon, S.A. Przybelski, B. Rawal, J.E. Parisi, R.C. Petersen, K. Kantarci, O.A. Ross, R. Duara, D.S. Knopman, C.R. Jack Jr, D.W. Dickson, Clinicopathologic and 11C-Pittsburgh compound B implications of Thal amyloid phase across the Alzheimer’s disease spectrum, Brain. 138 (2015) 1370–1381.

14. S.B. Wharton, T. Minett, D. Drew, G. Forster, F. Matthews, C. Brayne, P.G. Ince, on behalf of the MRC Cognitive Function and Ageing Neuropathology Study Group, Epidemiological pathology of Tau in the ageing brain: application of staging for neuropil threads (BrainNet Europe protocol) to the MRC cognitive function and ageing brain study, Acta Neuropathologica Communications. 4 (2016) 11.

15. S.B. Wharton, D. Wang, C. Parikh, F.E. Matthews, C. Brayne, P.G. Ince, Epidemiological pathology of Aβ deposition in the ageing brain in CFAS: addition of multiple Aβ-derived measures does not improve dementia assessment using logistic regression and machine learning approaches, Acta Neuropathologica Communications. 7 (2019) 1–12.

16. G. Lace, G.M. Savva, G. Forster, R. de Silva, C. Brayne, F.E. Matthews, J.J. Barclay, L. Dakin, P.G. Ince, S.B. Wharton, MRC-CFAS, Hippocampal tau pathology is related to neuroanatomical connections: an ageing population-based study, Brain. 132 (2009) 1324–1334.

17. P.G. Ince, T. Minett, G. Forster, C. Brayne, S.B. Wharton, M.R.C.C. Function, A.N. Study, Microinfarcts in an older population-representative brain donor cohort (MRC CFAS): Prevalence, relation to dementia and mobility, and implications for the evaluation of cerebral Small Vessel Disease, Neuropathol. Appl. Neurobiol. 43 (2017) 409–418.

18. H. Braak, E. Braak, Neuropathological stageing of Alzheimer-related changes, Acta Neuropathol. 82 (1991) 239–259.

19. H. Braak, I. Alafuzoff, T. Arzberger, H. Kretzschmar, K. Del Tredici, Staging of Alzheimer disease-associated neurofibrillary pathology using paraffin sections and immunocytochemistry, Acta Neuropathol. 112 (2006) 389–404.

20. I. Alafuzoff, D.R. Thal, T. Arzberger, N. Bogdanovic, S. Al-Sarraj, I. Bodi, S. Boluda, O. Bugiani, C. Duyckaerts, E. Gelpi, S. Gentleman, G. Giaccone, M. Graeber, T. Hortobagyi, R. Höftberger, P. Ince, J.W. Ironside, N. Kavantzas, A. King, P. Korkolopoulou, G.G. Kovács, D. Meyronet, C. Monoranu, T. Nilsson, P. Parchi, E. Patsouris, M. Pikkarainen, T. Revesz, A. Rozemuller, D. Seilhean, W. Schulz-Schaeffer, N. Streichenberger, S.B. Wharton, H. Kretzschmar, Assessment of β-amyloid deposits in human brain: a study of the BrainNet Europe Consortium, Acta Neuropathologica. 117 (2009) 309–320. https://doi.org/10.1007/s00401-009-0485-4.

21. D.R. Thal, U. Rüb, M. Orantes, H. Braak, Phases of Aβ-deposition in the human brain and its relevance for the development of AD, Neurology. 58 (2002) 1791–1800. https://doi.org/10.1212/wnl.58.12.1791.

22. J.F. Crary, J.Q. Trojanowski, J.A. Schneider, J.F. Abisambra, E.L. Abner, I. Alafuzoff, S.E. Arnold, J. Attems, T.G. Beach, E.H. Bigio, N.J. Cairns, D.W. Dickson, M. Gearing, L.T. Grinberg, P.R. Hof, B.T. Hyman, K. Jellinger, G.A. Jicha, G.G. Kovacs, D.S. Knopman, J. Kofler, W.A. Kukull, I.R. Mackenzie, E. Masliah, A. McKee, T.J. Montine, M.E. Murray, J.H. Neltner, I. Santa-Maria, W.W. Seeley, A. Serrano-Pozo, M.L. Shelanski, T. Stein, M. Takao, D.R. Thal, J.B. Toledo, J.C. Troncoso, J.P. Vonsattel, C.L. White 3rd, T. Wisniewski, R.L. Woltjer, M. Yamada, P.T. Nelson, Primary age-related tauopathy (PART): a common pathology associated with human aging, Acta Neuropathol. 128 (2014) 755–766.

23. S.B. Wharton, on behalf of the Cognitive Function and Ageing Neuropathology Study Group, D. Wang, C. Parikh, F.E. Matthews, C. Brayne, P.G. Ince, Epidemiological pathology of Aβ deposition in the ageing brain in CFAS: addition of multiple Aβ-derived measures does not improve dementia assessment using logistic regression and machine learning approaches, Acta Neuropathologica Communications. 7 (2019). https://doi.org/10.1186/s40478-019-0858-4.

24. S. Love, K. Chalmers, P. Ince, M. Esiri, J. Attems, K. Jellinger, M. Yamada, M. McCarron, T. Minett, F. Matthews, S. Greenberg, D. Mann, P.G. Kehoe, Development, appraisal, validation and implementation of a consensus protocol for the assessment of cerebral amyloid angiopathy in post-mortem brain tissue, Am. J. Neurodegener. Dis. 3 (2014) 19–32.

25. K. Ikeda, Glial fibrillary tangles and argyrophilic threads: Classification and disease specificity, Neuropathology. 16 (1996) 71–77. https://doi.org/10.1111/j.1440-1789.1996.tb00158.x.

26. K. Ikeda, H. Akiyama, T. Arai, T. Nishimura, Glial Tau Pathology in Neurodegenerative Diseases: Their Nature and Comparison with Neuronal Tangles, Neurobiology of Aging. 19 (1998) S85–S91. https://doi.org/10.1016/s0197-4580(98)00034-7.

27. K. Ikeda, H. Akiyama, H. Kondo, C. Haga, E. Tanno, T. Tokuda, S. Ikeda, Thorn-shaped astrocytes: possibly secondarily induced tau-positive glial fibrillary tangles, Acta Neuropathologica. 90 (1995) 620–625. https://doi.org/10.1007/bf00318575.

28. M. Nishimura, Y. Namba, K. Ikeda, M. Oda, Glial fibrillary tangles with straight tubules in the brains of patients with progressive supranuclear palsy, Neuroscience Letters. 143 (1992) 35–38. https://doi.org/10.1016/0304-3940(92)90227-x.

29. R.E. Marioni, F.E. Matthews, C. Brayne, MRC Cognitive Function and Ageing Study, The association between late-life cognitive test scores and retrospective informant interview data, Int. Psychogeriatr. 23 (2011) 274–279.

30. Huan Liu, R. Setiono, Chi2: feature selection and discretization of numeric attributes, in: Proceedings of 7th IEEE International Conference on Tools with Artificial Intelligence, 1995: pp. 388–391.

31. I. Kononenko, On biases in estimating multi-valued attributes, in: Ijcai, Citeseer, 1995: pp. 1034–1040.

32. J.R. Quinlan, Induction of decision trees, Machine Learning. 1 (1986) 81–106. https://doi.org/10.1007/bf00116251.

33. M. Robnik-Šikonja, I. Kononenko, Machine Learning, 53 (2003) 23–69. https://doi.org/10.1023/a:1025667309714.

34. J. Novakovic, P. Strbac, D. Bulatovic, Toward optimal feature selection using ranking methods and classification algorithms, Yugoslav Journal of Operations Research. 21 (2011) 119–135. https://doi.org/10.2298/yjor1101119n.

35. L. Yu, H. Liu, Efficient Feature Selection via Analysis of Relevance and Redundancy, J. Mach. Learn. Res. 5 (2004) 1205–1224.

36. F. Thabtah, F. Kamalov, S. Hammoud, S.R. Shahamiri, Least Loss: A simplified filter method for feature selection, Inf. Sci. . 534 (2020) 1–15.

37. K.D. Rajab, New Hybrid Features Selection Method: A Case Study on Websites Phishing, Security and Communication Networks. 2017 (2017). https://doi.org/10.1155/2017/9838169.

38. F. Kamalov, F. Thabtah, A Feature Selection Method Based on Ranked Vector Scores of Features for Classification, Annals of Data Science. 4 (2017) 483–502. https://doi.org/10.1007/s40745-017-0116-1.

39. M. Rajab, D. Wang, Practical Challenges and Recommendations of Filter Methods for Feature Selection, J. Info. Know. Mgmt. (2020) 2040019.

40. M.A. Hall, Correlation-based Feature Selection for Machine Learning, 1999.

41. M. Hall, E. Frank, G. Holmes, B. Pfahringer, P. Reutemann, I.H. Witten, The WEKA data mining software: an update, ACM SIGKDD Explorations Newsletter. 11 (2009) 10–18.

42. F. Pedregosa, G. Varoquaux, A. Gramfort, V. Michel, B. Thirion, O. Grisel, M. Blondel, P. Prettenhofer, R. Weiss, V. Dubourg, Others, Scikit-learn: Machine learning in Python, The Journal of Machine Learning Research. 12 (2011) 2825–2830.

43. X. Lin, C. Li, Y. Zhang, B. Su, M. Fan, H. Wei, Selecting Feature Subsets Based on SVM-RFE and the Overlapping Ratio with Applications in Bioinformatics, Molecules. 23 (2017). https://doi.org/10.3390/molecules23010052.

44. J. Xia, L. Sun, S. Xu, Q. Xiang, J. Zhao, W. Xiong, Y. Xu, S. Chu, A Model Using Support Vector Machines Recursive Feature Elimination (SVM-RFE) Algorithm to Classify Whether COPD Patients Have Been Continuously Managed According to GOLD Guidelines, International Journal of Chronic Obstructive Pulmonary Disease. 15 (2020) 2779–2786. https://doi.org/10.2147/copd.s271237.

45. Y. Saeys, I. Inza, P. Larrañaga, A review of feature selection techniques in bioinformatics, Bioinformatics. 23 (2007) 2507–2517.

46. T. Chen, C. Guestrin, XGBoost, Proceedings of the 22nd ACM SIGKDD International Conference on Knowledge Discovery and Data Mining. (2016). https://doi.org/10.1145/2939672.2939785.

47. I. Alafuzoff, T. Arzberger, S. Al-Sarraj, I. Bodi, N. Bogdanovic, H. Braak, O. Bugiani, K. Del-Tredici, I. Ferrer, E. Gelpi, G. Giaccone, M.B. Graeber, P. Ince, W. Kamphorst, A. King, P. Korkolopoulou, G.G. Kovács, S. Larionov, D. Meyronet, C. Monoranu, P. Parchi, E. Patsouris, W. Roggendorf, D. Seilhean, F. Tagliavini, C. Stadelmann, N. Streichenberger, D.R. Thal, S.B. Wharton, H. Kretzschmar, Staging of neurofibrillary pathology in Alzheimer’s disease: a study of the BrainNet Europe Consortium, Brain Pathol. 18 (2008) 484–496.

48. A. Serrano-Pozo, J. Qian, S.E. Monsell, D. Blacker, T. Gómez-Isla, R.A. Betensky, J.H. Growdon, K.A. Johnson, M.P. Frosch, R.A. Sperling, B.T. Hyman, Mild to moderate Alzheimer dementia with insufficient neuropathological changes, Ann. Neurol. 75 (2014) 597–601.

49. H.A.D. Keage, P.G. Ince, F.E. Matthews, S.B. Wharton, I.G. McKeith, C. Brayne, on behalf of MRC CFAS and CC75C, Impact of less common and “disregarded” neurodegenerative pathologies on dementia burden in a population-based cohort, J. Alzheimers. Dis. 28 (2012) 485–493.

50. M. M. Corrada, D. J. Berlau, C. H. Kawas, A Population-Based Clinicopathological Study in the Oldest-Old: The 90+ Study, Curr. Alzheimer Res. 9 (2012) 709–717.

51. A. Hall, T. Pekkala, T. Polvikoski, M. van Gils, M. Kivipelto, J. Lötjönen, J. Mattila, M. Kero, L. Myllykangas, M. Mäkelä, M. Oinas, A. Paetau, H. Soininen, M. Tanskanen, A. Solomon, Prediction models for dementia and neuropathology in the oldest old: the Vantaa 85+ cohort study, Alzheimers. Res. Ther. 11 (2019) 11.

52. I. Jain, V.K. Jain, R. Jain, Correlation feature selection based improved-Binary Particle Swarm Optimization for gene selection and cancer classification, Applied Soft Computing. 62 (2018) 203–215. https://doi.org/10.1016/j.asoc.2017.09.038.

53. M.W. Mwadulo, A review on feature selection methods for classification tasks, (2016). http://citeseerx.ist.psu.edu/viewdoc/download?doi=10.1.1.1075.7828&rep=rep1&type=pdf (accessed April 6, 2021).

54. H. Shi, H. Li, D. Zhang, C. Cheng, X. Cao, An efficient feature generation approach based on deep learning and feature selection techniques for traffic classification, Computer Networks. 132 (2018) 81–98.

55. W. Gómez Flores, W.C. de A. Pereira, A.F.C. Infantosi, Improving classification performance of breast lesions on ultrasonography, Pattern Recognit. 48 (2015) 1125–1136.

56. B. Agarwal, N. Mittal, Prominent feature extraction for review analysis: an empirical study, J. Exp. Theor. Artif. Intell. 28 (2016) 485–498.

57. J. Izetta, P.F. Verdes, P.M. Granitto, Improved multiclass feature selection via list combination, Expert Syst. Appl. 88 (2017) 205–216.

58. O. Stromann, A. Nascetti, O. Yousif, Y. Ban, Dimensionality Reduction and Feature Selection for Object-Based Land Cover Classification based on Sentinel-1 and Sentinel-2 Time Series Using Google Earth Engine, Remote Sensing. 12 (2019) 76.

59. J. Gonzalez-Lopez, S. Ventura, A. Cano, Distributed multi-label feature selection using individual mutual information measures, Knowledge-Based Systems. 188 (2020) 105052.

60. G. Lace, P.G. Ince, C. Brayne, G.M. Savva, F.E. Matthews, R. de Silva, J.E. Simpson, S.B. Wharton, Mesial temporal astrocyte tau pathology in the MRC-CFAS ageing brain cohort, Dement. Geriatr. Cogn. Disord. 34 (2012) 15–24.

61. A. Keo, A. Mahfouz, A.M.T. Ingrassia, J.-P. Meneboo, C. Villenet, E. Mutez, T. Comptdaer, B.P.F. Lelieveldt, M. Figeac, M.-C. Chartier-Harlin, W.D.J. van de Berg, J.J. van Hilten, M.J.T. Reinders, Transcriptomic signatures of brain regional vulnerability to Parkinson’s disease, Commun Biol. 3 (2020) 101.

62. M.W. Weiner, P.S. Aisen, C.R. Jack Jr, W.J. Jagust, J.Q. Trojanowski, L. Shaw, A.J. Saykin, J.C. Morris, N. Cairns, L.A. Beckett, A. Toga, R. Green, S. Walter, H. Soares, P. Snyder, E. Siemers, W. Potter, P.E. Cole, M. Schmidt, Alzheimer’s Disease Neuroimaging Initiative, The Alzheimer’s disease neuroimaging initiative: progress report and future plans, Alzheimers. Dement. 6 (2010) 202–11.e7.

63. D.A. Bennett, J.A. Schneider, A.S. Buchman, C. Mendes de Leon, J.L. Bienias, R.S. Wilson, The Rush Memory and Aging Project: study design and baseline characteristics of the study cohort, Neuroepidemiology. 25 (2005) 163–175.

64. F.M. Elahi, B.L. Miller, A clinicopathological approach to the diagnosis of dementia, Nature Reviews Neurology. 13 (2017) 457–476. https://doi.org/10.1038/nrneurol.2017.96.

65. W.W. Barker, C.A. Luis, A. Kashuba, M. Luis, D.G. Harwood, D. Loewenstein, C. Waters, P. Jimison, E. Shepherd, S. Sevush, N. Graff-Radford, D. Newland, M. Todd, B. Miller, M. Gold, K. Heilman, L. Doty, I. Goodman, B. Robinson, G. Pearl, D. Dickson, R. Duara, Relative frequencies of Alzheimer disease, Lewy body, vascular and frontotemporal dementia, and hippocampal sclerosis in the State of Florida Brain Bank, Alzheimer Dis. Assoc. Disord. 16 (2002) 203–212.

66. D.S. Geldmacher, P.J. Whitehouse, Evaluation of dementia, N. Engl. J. Med. 335 (1996) 330–336.

67. A. Hoque, S. Galib, M. Tasnim, Mining pathological data to support medical diagnostics, in: Workshop on Advances on Data Management: Applications and Algorithms, Department of Computer Science and Engineering, BUET, Dhaka, academia.edu, 2013: pp. 71–74.

68. F. Kherif, S. Muller, Neuro-Clinical Signatures of Language Impairments: A Theoretical Framework for Function-to-structure Mapping in Clinics, Curr. Top. Med. Chem. 20 (2020) 800–811.

69. T.A. Allen, A.M. Schreiber, N.T. Hall, M.N. Hallquist, From Description to Explanation: Integrating Across Multiple Levels of Analysis to Inform Neuroscientific Accounts of Dimensional Personality Pathology, J. Pers. Disord. 34 (2020) 650–676.

70. C. Gaiteri, S. Mostafavi, C.J. Honey, P.L. De Jager, Genetic variants in Alzheimer disease— molecular and brain network approaches, Nat. Rev. (2016). https://www.nature.com/articles/nrneurol.2016.84.pdf?origin=ppub.

71. X. Zhou, S. Chen, B. Liu, R. Zhang, Y. Wang, P. Li, Y. Guo, H. Zhang, Z. Gao, X. Yan, Development of traditional Chinese medicine clinical data warehouse for medical knowledge discovery and decision support, Artif. Intell. Med. 48 (2010) 139–152.

