## Supplementary Table 1 for "Assessment of Alzheimer-related Pathologies of Dementia Using Machine Learning Feature Selection"

### Supplementary Information

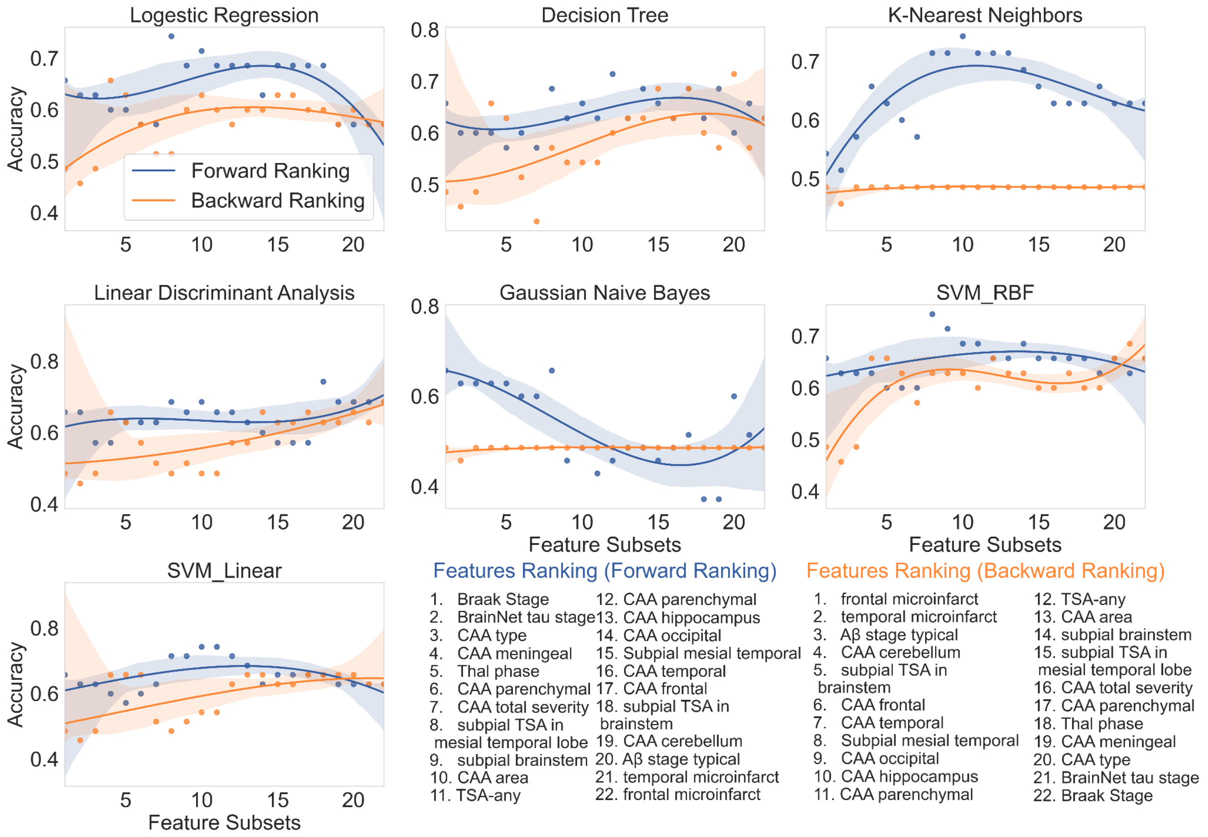


Figure 1: *Accuracy performance of all subsets of neuropathology features from the rank list forward and backward rankings. Forward ranking (blue) adds to the classifier model from the top feature to the lowest feature while the backward ranking (orange) adds to the model from the lowest feature to the top feature. Seven classifiers were utilized in this investigation: Logistic Regression, Decision Tree, k-Nearest Neighbors, Linear Discriminant Analysis, Gaussian Naive Bayes , Support Vector Machines with Radial Basis Function kernel, and Support Vector Machines with Linear kernel.*

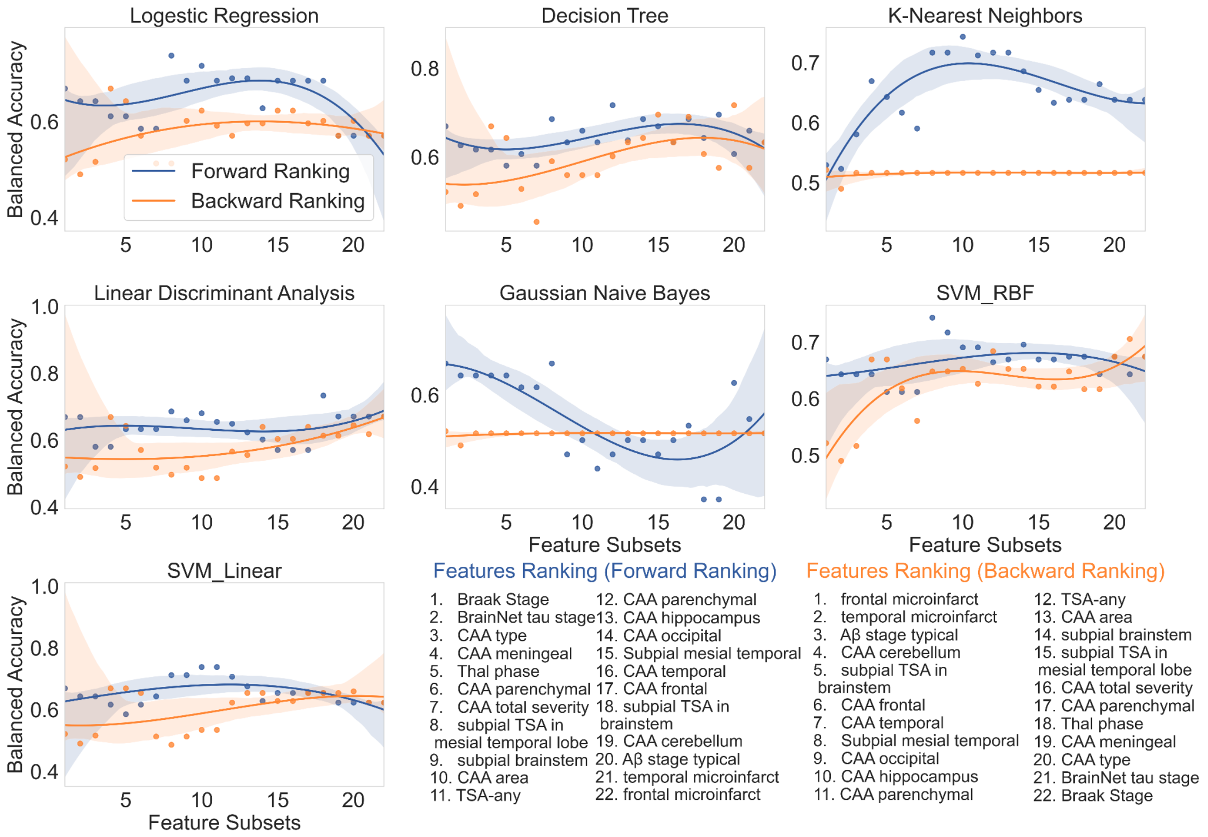


Figure 2: *Balanced Accuracy performance of all subsets of neuropathology features from the rank list forward and backward rankings. Forward ranking (blue) adds to the classifier model from the top feature to the lowest feature while the backward ranking (orange) adds to the model from the lowest feature to the top feature. Seven classifiers were utilized in this investigation: Logistic Regression, Decision Tree, k-Nearest Neighbors, Linear Discriminant Analysis, Gaussian Naive Bayes , Support Vector Machines with Radial Basis Function kernel, and Support Vector Machines with Linear kernel.*

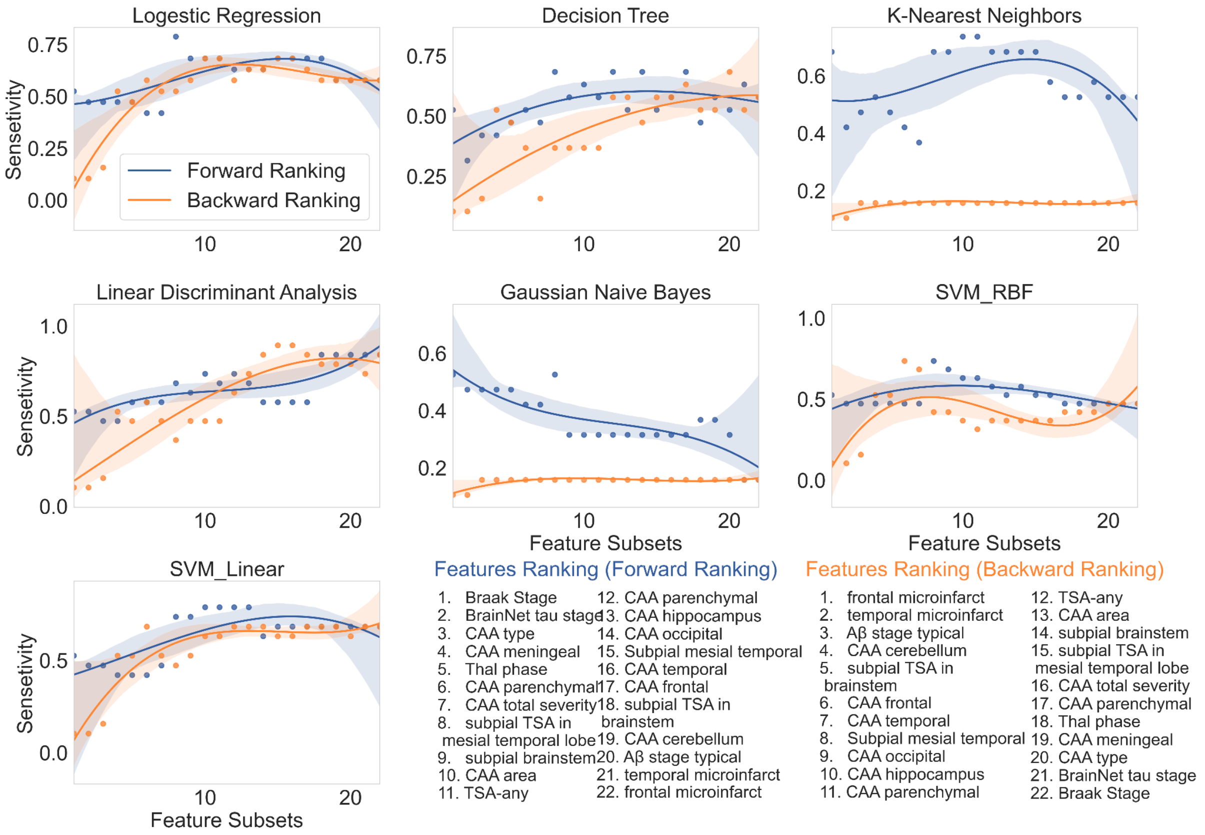


Figure 3: *Sensitivity performance of all subsets of neuropathology features from the rank list forward and backward rankings. Forward ranking (blue) adds to the classifier model from the top feature to the lowest feature while the backward ranking (orange) adds to the model from the lowest feature to the top feature. Seven classifiers were utilized in this investigation: Logistic Regression, Decision Tree, k-Nearest Neighbors, Linear Discriminant Analysis, Gaussian Naive Bayes , Support Vector Machines with Radial Basis Function kernel, and Support Vector Machines with Linear kernel.*

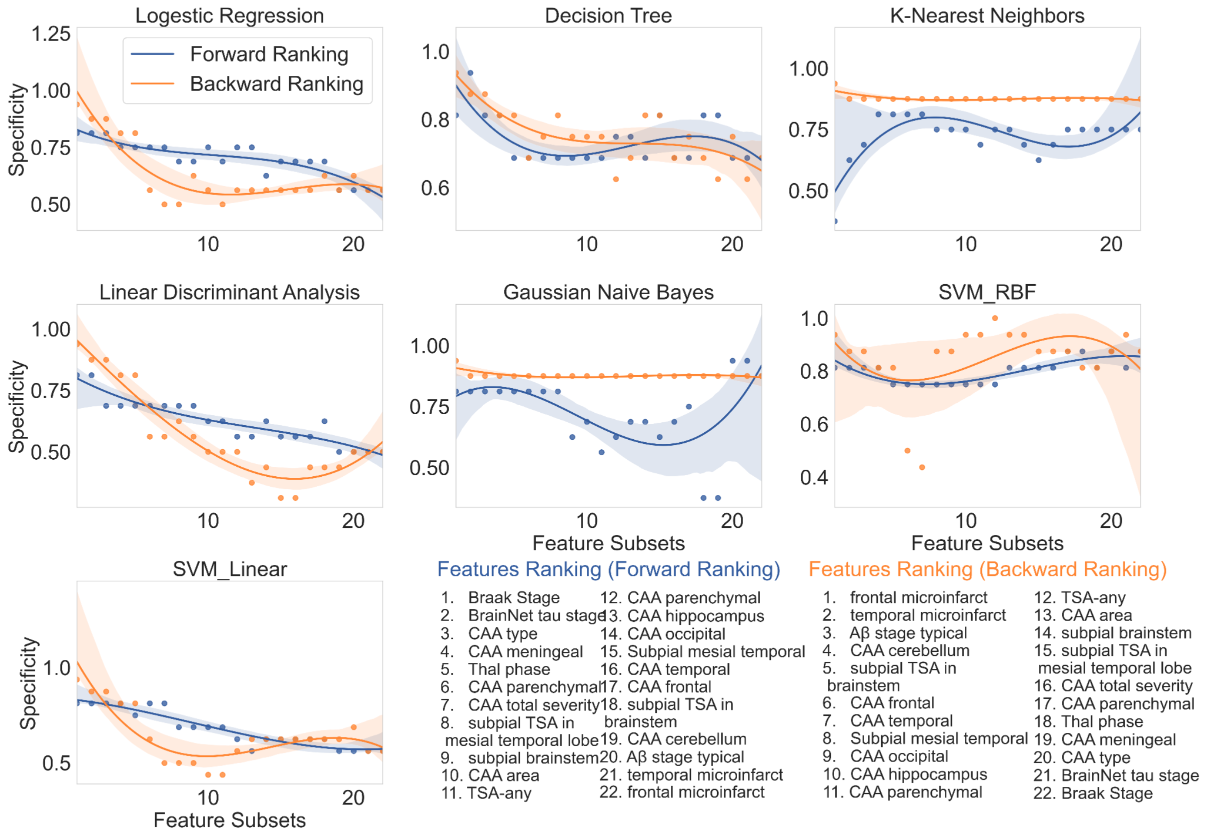


Figure 4: *Specificity performance of all subsets of neuropathology features from the rank list forward and backward rankings. Forward ranking (blue) adds to the classifier model from the top feature to the lowest feature while the backward ranking (orange) adds to the model from the lowest feature to the top feature. Seven classifiers were utilized in this investigation: Logistic Regression, Decision Tree, k-Nearest Neighbors, Linear Discriminant Analysis, Gaussian Naive Bayes , Support Vector Machines with Radial Basis Function kernel, and Support Vector Machines with Linear kernel.*

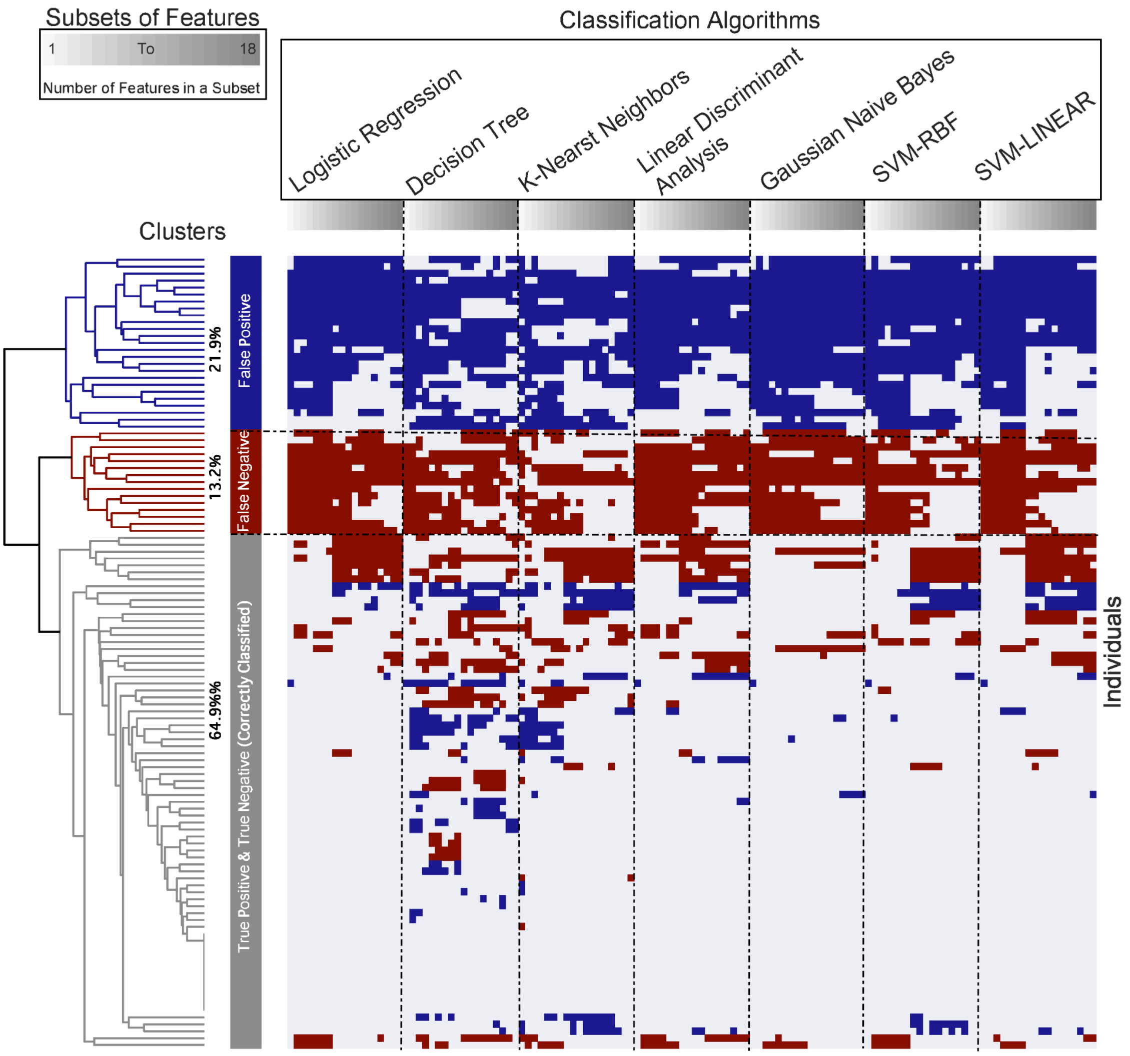


Figure 5: *Clustering of cross-validation classification performance on 114 CFAS participants and subsets of 18 features including 8 top ranked neuropathology features and 10 non-standard neuropathology features. Each cluster illustrates a classification that was given to individuals consistently, or nearly consistently, irrespective of what classification algorithm was used. Evaluation of 7 classifiers revealed 23 individuals (blue) were mostly misclassified as false positive, 15 individuals (red) were mostly misclassified as false negative, and 76 individuals (grey) were mostly correctly classified as true positive or true negative.*

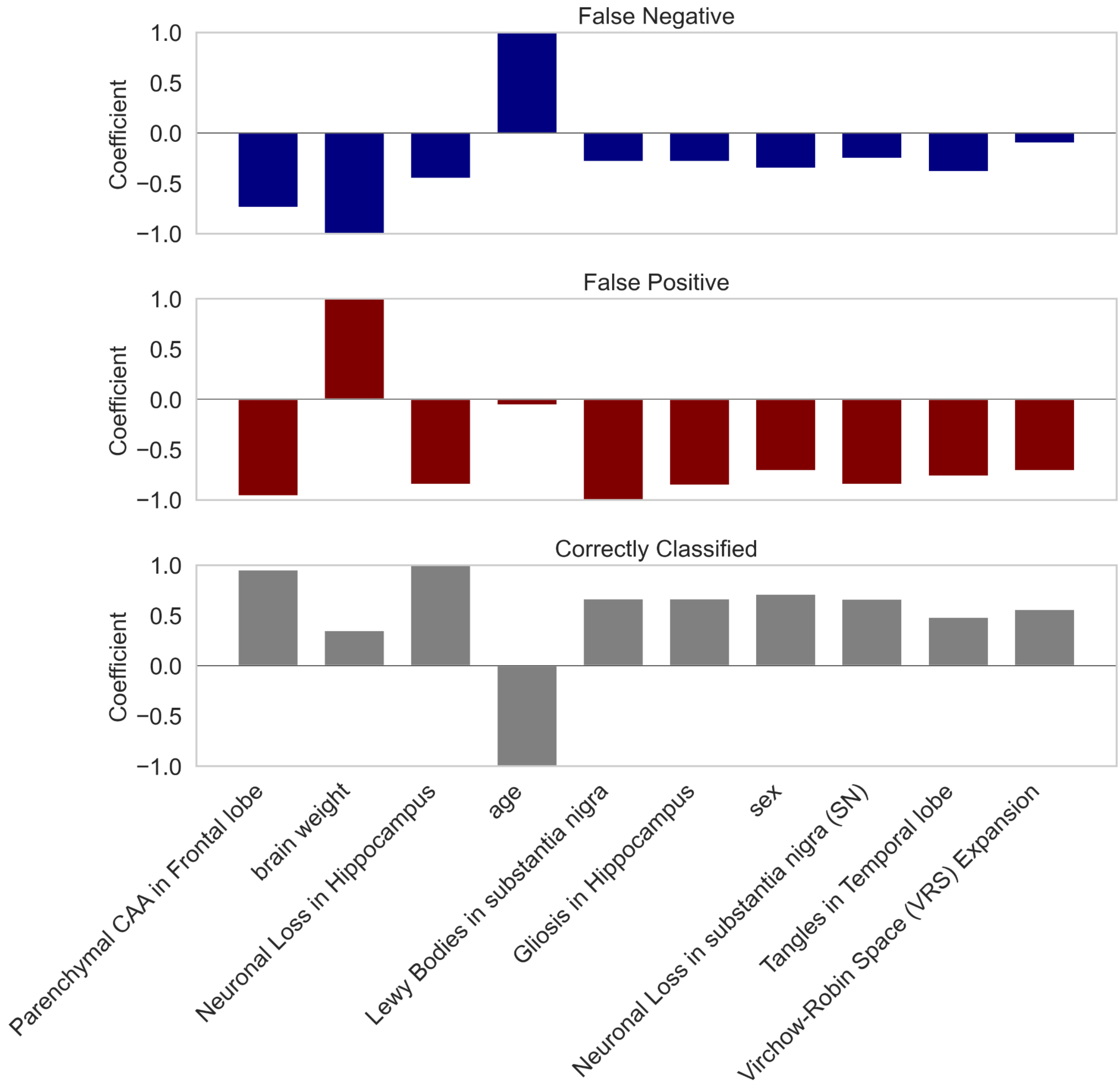


Figure 6: *Non-standard neuropathological and demographic features that were associated with mis-classified and correctly classified cases by the classic neuropathology features. The coefficients shown for each variable were extracted from the most predictive support vector machine classifiers: demographics features such as, age, brain weight and sex.*

Table 1: *Ranking of the CFAS Dataset Features using Different Feature Selection Techniques. Seven feature-ranking methods were presented: Chi-Square (CHI), Gain Ratio (GR), Information Gain (IG), ReliefF (RF), Symmetrical Uncertainty (SmyUn), Least Loss (L2) and Variable Analysis (Va).*

| NO | Chi-Squares | Gain Ratio | Information Gain | ReliefF | Symmetrical Uncertainty | Least Loss | Variable Analysis |
|---|---|---|---|---|---|---|---|
| **1** | BraakStage | age | BraakStage | BrainNetStage | age | CAAType | BraakStage |
| **2** | BrainNetStage | CAATotalSev | BrainNetStage | BraakStage | BraakStage | brain weight | BrainNetStage |
| **3** | CAAMeningeal | AbStageTypical | CAAMeningeal | MTSPETSA | BrainNetStage | age | CAAMeningeal |
| **4** | CAAParenc | brain weight | CAAParenc | TSAAny | CAATotalSev | BrainNetStage | CAAParenc |
| **5** | age | BraakStage | age | SubpialMesTemp | brain weight | MTSPETSA | age |

| | | | | | | | |
|---|---|---|---|---|---|---|---|
| **6** | ThalStage | BrainNetStage | ThalStage | ThalStage | CAAType | BraakStage | ThalStage |
| **7** | CAAType | CAAAreas | CAAType | age | CAAMeningeal | CAAAreas | CAAType |
| **8** | brain weight | SubpialBrainstem | brain weight | CAAType | CAAParenc | CAAHippocampus | brain weight |
| **9** | SubpialBrainstem | CAAType | CAATotalSev | TSATotal | SubpialBrainstem | SubpialBrainstem | SubpialBrainste m |
| **10** | CAAAreas | MTSPETSA | SubpialBrainstem | SubpialBrainstem | CAAAreas | CAAParietal | CAATotalSev |
| **11** | CAATotalSev | BSSPETSA | CAAAreas | HippocTauStage | ThalStage | ThalStage | CAAAreas |
| **12** | MTSPETSA | CAAMeningeal | MTSPETSA | SubcorticalStage | MTSPETSA | CAAFrontal | MTSPETSA |
| **13** | CAAHippocampus | TempMicroinf | CAAHippocampus | brain weight | BSSPETSA | CAATemp | CAAHippocamp us |
| **14** | CAAParietal | CAAParenc | CAAParietal | BSSPETSA | CAAHippocampus | CAAOccipital | CAAParietal |
| **15** | CAAFrontal | ThalStage | CAAFrontal | CAAOccipital | CAAParietal | TSAAny | CAAFrontal |
| **16** | CAAOccipital | CAAHippocampus | BSSPETSA | CAAParietal | CAAOccipital | CAACerebellum | CAAOccipital |
| **17** | HippocTauStage | CAAOccipital | CAAOccipital | PARTall | CAAFrontal | CAAMeningeal | HippocTauStage |
| **18** | CAATemp | CAAParietal | HippocTauStage | CAAMeningeal | CAATemp | SubpialMesTemp | BSSPETSA |
| **19** | BSSPETSA | CAAFrontal | CAATemp | CAAHippocampus | TSAAny | CAAParenc | CAATemp |
| **20** | TSAAny | CAATemp | TSAAny | MicroinfarctStage | CAACerebellum | CAATotalSev | TSAAny |
| **21** | CAACerebellum | TSAAny | CAACerebellum | CAACerebellum | AbStageTypical | HippocTauStage | CAACerebellum |
| **22** | SubpialMesTemp | FrontalMicroin | SubpialMesTemp | CAAParenc | SubpialMesTemp | BSSPETSA | SubpialMesTem p |
| **23** | AbStageTypical | CAACerebellum | AbStageTypical | CAATotalSev | HippocTauStage | FrontalMicroin | AbStageTypical |
| **24** | TempMicroinf | SubpialMesTemp | TempMicroinf | CAAAreas | TempMicroinf | TempMicroinf | TempMicroinf |
| **25** | FrontalMicroin | HippocTauStage | FrontalMicroin | CxSPETSA | FrontalMicroin | PARTdefinite | FrontalMicroin |
| **26** | PARTdefinite | ArgyrGrains | PARTdefinite | PARTdefinite | ParMicrin | AbStageTypical | PARTdefinite |
| **27** | ParMicrin | ParMicrin | ParMicrin | ArgyrGrains | PARTdefinite | ParMicrin | ParMicrin |
| **28** | ArgyrGrains | PARTdefinite | ArgyrGrains | TempMicroinf | ArgyrGrains | OccipMicroing | ArgyrGrains |
| **29** | OccipMicroing | CxSPETSA | OccipMicroing | CorticalStage | OccipMicroing | ArgyrGrains | OccipMicroing |
| **30** | CxSPETSA | OccipMicroing | CxSPETSA | AbStageTypical | CxSPETSA | PARTall | CxSPETSA |
| **31** | TuftedAst | TuftedAst | TuftedAst | FrontalMicroin | TuftedAst | TuftedAst | TuftedAst |
| **32** | PARTall | PARTall | PARTall | OccipMicroing | PARTall | CxSPETSA | PARTall |
| **33** | MicroinfarctStage | CorticalStage | SubcorticalStage | CAATemp | SubcorticalStage | CorticalStage | MicroinfarctStae |
| **34** | TSATotal | TSATotal | MicroinfarctStage | ParMicrin | CorticalStage | SubcorticalStage | TSATotal |
| **35** | SubcorticalStage | SubcorticalStage | TSATotal | CAAFrontal | TSATotal | TSATotal | SubcorticalStage |
| **36** | CorticalStage | MicroinfarctStage | CorticalStage | TuftedAst | MicroinfarctStage | MicroinfarctStage | CorticalStage |

### Glossary List

| **Feature** | **Description** |
|---|---|
| **BraakStage** | Refers to Braak Neurofibrillary Tangle Stage (0-VI) (Braak, Alafuzoff et al. 2006). Braak stages: (I/II) when neurofibrillary tangle involvement is limited to the transentorhinal region of the brain, stages |

| | |
|---|---|
| | (III/IV) when involvement of limbic regions such as the hippocampus appears.<br>(V/VI) when there is extensive neocortical involvement. |
| **BrainNetStage** | Brain-Net Tau Stage (1-6) - (Brain-Net Europe protocol for tau pathology) |
| **Aged** | Age of an individual at death |
| **CAATotalSev** | Cerebral amyloid angiopathy (CAA) total severity is the scores for leptomeningeal and parenchymal amyloid, giving a score out of maximum of 24 for severity in cortical areas. |
| **CAAMeningeal** | Cerebral amyloid angiopathy (CAA) Severity Meningeal as for parenchymal the scores are out of 12 |
| **Brain-weight** | Brain weight of an individual after death |
| **CAAType:** | Cerebral amyloid angiopathy (CAA) type as defined by Thal, (1) are cases with capillary amyloid and (2) only in larger vessels, (0) no CAA |
| **ThalStage** | Thal Abeta stage- the new brain -Net-Stage for Abeta. This is a five-stage scheme. This is based on the detection of immunopositive amyloid in cortical and subcortical areas,<br>(1) progressive deposition of amyloid in neocortex,<br>(2) allocortex or limbic,<br>(3) diencephalon/basal ganglia<br>(4) brainstem/midbrain,<br>(5) cerebellum |
| **MTSPETSA** | Thorn-Shaped Astrcytes (TSA) subpial/epen mesial temporal |
| **CAAAreas** | Number of brain areas examined that have cerebral amyloid angiopathy (CAA). The number of anatomical areas involved from all the areas in the sampling set to obtain a measure of the extent (number of areas out of 9 maximum). |
| **Subpialbrainstem** | Subpial tau neurites in brainstem/subcortical region |
| **CAAParenc** | Cerebral amyloid angiopathy (CAA) Severity score (Love 2014) Parenchymal-So in any area CAA can be 1, 2 or 3 |

| | |
|---|---|
| **TSATotal** | Thorn-Shaped Astrcytes (TSA), presence (1) or absence (0) TSA in any brain area |
| **TSAAny** | Thorn-Shaped Astrcytes (TSA), Total number of areas with TSA |
| **CAAParietal** | Cerebral amyloid angiopathy (CAA) is present or not in Parietal Cortex |
| **CAAHippocampus** | Cerebral amyloid angiopathy (CAA) is present or not in Hippocampus and (OTG) occipitotemporal gyrus |
| **CAAFrontal** | Cerebral amyloid angiopathy (CAA) is present or not in Frontal Cortex |
| **CAAOcciptal** | Cerebral amyloid angiopathy (CAA) is present or not in Occipital Cortex |
| **BSSPETSA** | Thorn-Shaped Astrcytes (TSA) subpial/epend brainstem |
| **CAACerebellum** | Cerebral amyloid angiopathy (CAA) is present or not in Cerebellum |
| **AbStageTypical** | (1) Thal typical, (2) atypical |
| **HippocTauStage** | Hippocampal Lace Tau Stage based on progression through the hippocampus |
| **SubcorticalStage** | Subcortical lacune stage- number of subcortical areas that have microinfarcts |
| **CorticalStage** | Cortical microinfarct stage- number of cortical areas that have microinfarcts |
| **SubpialMesTemp** | Subpial tau neurites in mesial temporal |
| **PARTall** | Primary age-related tauopathy (PART), (1) PARTall Abeta 0-2/tau I-IV, (2) Abeta 3+/tau I-IV |
| **MicroinfractStage** | Total microinfarct stage- number of total of areas that have microinfarcts |
| **ArgyGrains** | Argyrophilic grains |

| | |
|---|---|
| **OccipMicroing** | Occipital Microinfarct |
| **ParMicrin** | Parietal Microinfarct |
| **PARTdefinite** | Primary age-related tauopathy (PART), PART-definite is defined as no Aβ pathology (Thal stage 0) and Braak NFT stage I-IV and PART-possible as mild Aβ pathology (Thal stage I-II)/Braak NFT stage I-IV (Crary, Trojanowski et al. 2014). |
| **FrontalMicroin** | Frontal Microinfarct |
| **TempMicroinf** | Temporal Microinfarct |
| **CxSPETSA** | Thorn-Shaped Astrcytes (TSA) subpial/epend cortex |
| **CAATemp** | Cerebral amyloid angiopathy (CAA) is present or not in Temporal Cortex |
| **TuffedAst** | Tufted parencymal astrocytes any area |
